# A Learning Health System Randomized Trial of Monoclonal Antibodies for Covid-19

**DOI:** 10.1101/2021.09.03.21262551

**Authors:** Erin K. McCreary, J. Ryan Bariola, Tami Minnier, Richard J. Wadas, Judith A. Shovel, Debbie Albin, Oscar C. Marroquin, Kevin E. Kip, Kevin Collins, Mark Schmidhofer, Mary Kay Wisniewski, David A Nace, Colleen Sullivan, Meredith Axe, Russell Meyer, Alexandra Weissman, William Garrard, Octavia M. Peck-Palmer, Alan Wells, Robert D. Bart, Anne Yang, Lindsay Berry, Scott Berry, Amy Crawford, Anna McGlothin, Tina Khadem, Kelsey Linstrum, Stephanie K. Montgomery, Daniel Ricketts, Jason N. Kennedy, Caroline J. Pidro, Ghady Haidar, Graham M. Snyder, Bryan J. McVerry, Derek C. Angus, Paula L. Kip, Christopher W. Seymour, David T. Huang

## Abstract

**Background:** Neutralizing monoclonal antibodies (mAb) targeting SARS-CoV-2 decrease hospitalization and death in patients with mild to moderate Covid-19. Yet, their clinical use is limited, and comparative effectiveness is unknown.

**Methods:** We present the first results of an ongoing, learning health system adaptive platform trial to expand mAb treatment to all eligible patients and evaluate the comparative effectiveness of available mAbs. The trial launched March 10, 2021. Results are reported as of June 25, 2021 due to the U.S. federal decision to pause distribution of bamlanivimab-etesevimab; patient follow-up concluded on July 23, 2021. Patients referred for mAb who met Emergency Use Authorization criteria were provided a random mAb allocation of bamlanivimab, bamlanivimab-etesevimab, or casirivimab-imdevimab with a therapeutic interchange policy. The primary outcome was hospital-free days (days alive and free of hospital) within 28 days, where patients who died were assigned -1 day. The primary analysis was a Bayesian cumulative logistic model of all patients treated at an infusion center or emergency department, adjusting for treatment location, age, sex, and time. Inferiority was defined as a 99% posterior probability of an odds ratio < 1. Equivalence was defined as a 95% posterior probability that the odds ratio is within a given bound.

**Results:** Prior to trial launch, 3.1% (502) of 16,345 patients who were potentially eligible by an automated electronic health record (EHR) screen received mAb. During the trial period, 23.2% (1,201) of 5,173 EHR-screen eligible patients were treated, a 7.5-fold increase. After including additional referred patients from outside the health system, a total of 1,935 study patients received mAb therapy (128 bamlanivimab, 885 bamlanivimab-etesevimab, 922 casirivimab-imdevimab). Mean age ranged from 55 to 57 years, half were female (range, 53% to 54%), and 17% were Black (range, 12% to 19%). Median hospital–free days were 28 (IQR, 28 to 28) for each mAb group. Hospitalization varied between groups (bamlanivimab, 12.5%; bamlanivimab-etesevimab, 14.7%, casirivimab-imdevimab, 14.3%). Relative to casirivimab-imdevimab, the median adjusted odds ratios were 0.58 (95% credible interval (CI), 0.30 to 1.16) and 0.94 (95% CI, 0.72 to 1.24) for the bamlanivimab and bamlanivimab-etesevimab groups, respectively. These odds ratios yielded 91% and 94% probabilities of inferiority of bamlanivimab versus bamlanivimab-etesevimab and casirivimab-imdevimab respectively, and an 86% probability of equivalence between bamlanivimab-etesevimab and casirivimab-imdevimab, at the prespecified odds ratio bound of 0.25. Twenty-one infusion-related adverse events occurred in 0% (0/128), 1.4% (12/885), and 1.0% (9/922) of patients treated with bamlanivimab, bamlanivimab-etesevimab, and casirivimab-imdevimab, respectively.

**Conclusion:** In non-hospitalized patients with mild to moderate Covid-19, bamlanivimab, compared to bamlanivimab-etesevimab and casirivimab-imdevimab, resulted in 91% and 94% probabilities of inferiority with regards to odds of improvement in hospital-free days within 28 days. There was an 86% probability of equivalence between bamlanivimab-etesevimab and casirivimab-imdevimab at an odds ratio bound of 0.25. However, the trial was unblinded early due to federal distribution decisions, and no mAb met prespecified criteria for statistical inferiority or equivalence. (ClinicalTrials.gov, NCT04790786).

## INTRODUCTION

Neutralizing monoclonal antibodies (mAb) targeting SARS-CoV-2 were granted U.S. Food and Drug Administration (FDA) Emergency Use Authorization (EUA) for treatment of mild to moderate Covid-19, and significantly decrease hospitalization and death in this patient population.^1-9^ However, clinical use is limited due to lack of access, logistical challenges of administration, and incomplete clinician and patient awareness.^10^ The comparative effectiveness of one mAb versus another is also unknown.^11^

In February 2021, UPMC partnered with the U.S. Federal Covid-19 Response Team to expand clinical use to all EUA-eligible patients and evaluate the comparative effectiveness of the available mAbs using a learning health system approach. This approach embeds knowledge generation into daily practice to seek continuous improvement in care.^12,13^ For all mAb-eligible patients treated at UPMC, we set two objectives, (i.) to equitably treat as broad of a proportion of mAb-eligible patients as possible, and (ii.) to compare effectiveness between mAbs overall and over time as SARS-CoV-2 variants emerged. This report presents the first results of the OPTIMISE-C19 (OPtimizing Treatment and Impact of Monoclonal antIbodieS through Evaluation for Covid-19) trial.^14^

## METHODS

### Expansion of Monoclonal Antibody Access

UPMC is an integrated network predominantly serving Western and Central Pennsylvania. Prior to U.S. federal partnership, UPMC developed a mAb treatment infrastructure in response to the bamlanivimab EUA issued November 9, 2020.^7^ The first patient was treated at a one of 18 outpatient infusion centers within this infrastructure on December 9, 2020.^15.16^ After U.S. federal partnership, mAb infusion capacity was expanded to all emergency departments (ED, n = 31), infusion center staffing and hours were increased, and a multifaceted outreach campaign to patients and clinicians was launched. UPMC partnered with government public health bodies, community outreach leaders, and neighboring health systems to increase awareness and referrals, and a home infusion company to provide treatment for homebound patients (Chartwell Pennsylvania, Oakdale, PA).

The UPMC Pharmacy and Therapeutics Committee developed a therapeutic interchange policy on November 21, 2020 in response to the issuance of an EUA for casirivimab-imdevimab.^9^ The policy considered all available mAb equivalent; a patient could receive any mAb based on local inventory. The policy was updated to include bamlanivimab-etesevimab on February 9, 2021 and to remove bamlanivimab on March 31, 2021 due to U.S. federal decisions.^8,17^ All pharmacies supplying all infusion sites had equal opportunity to order any available mAb from a central supply facility. All mAb were ordered by prescribers as a generic referral order and provided as per FDA EUA guidance (**Supplement**). Prescribers were required to provide and review all mAb EUA Fact Sheets with the patient at time of referral and explain the patient could receive any of the EUA-governed mAb.^7-9^

### Trial Design and Oversight

OPTIMISE-C19 is an open-label, pragmatic, comparative effectiveness, platform trial with response-adaptive randomization. The three mAb (bamlanivimab, bamlanivimab-etesevimab, and casirivimab-imdevimab) evaluated in this report were supplied by the U.S. federal government. The trial was approved by the UPMC Quality Improvement Committee and launched March 10, 2021 (Project ID 3280). The University of Pittsburgh Institutional Review Board considered provision of mAb therapy quality improvement and only the additional data collection and analyses represented research (STUDY21020179).

A custom application built into the electronic health record (EHR) linked local inventory to patient encounters and provided a random mAb allocation at the time of referral, within the therapeutic interchange policy.^18^ Allocation was initially assigned equally to available mAb. Patients provided verbal consent to receive mAb therapy as part of routine care. The prescriber and/or patient could request a specific mAb if desired.

### Patients

A centralized monoclonal antibody operations team confirmed patient eligibility upon referral. Patients were eligible for mAb if they met EUA criteria, which included patients with a positive SARS-CoV-2 polymerase chain reaction or antigen test, mild to moderate symptoms for 10 days or less, and risk factor(s) for progression to severe Covid-19. The EUA criteria excluded patients who required supplemental oxygen above baseline requirements, weighed < 40 kg, were < 12 years of age, or were hospitalized for Covid-19 (**Supplement**). Patients were screened in the ED or outpatient infusion centers.

### Outcomes

To evaluate clinical mAb use expansion, we screened for potential eligibility based on EHR identification of all outpatients with a positive SARS-CoV-2 polymerase chain reaction or antigen test performed within the health system and an EUA-defined risk factor for progression to severe disease. Race was derived from registration system data using fixed categories consistent with the Centers for Medicare & Medicaid Services EHR meaningful use dataset and the AMA Manual of Style.^19,20^ Pre-specified categories included non-Hispanic Black, non-Hispanic White, and Other. Individuals were considered Other due to small sample sizes for Hispanic, American Indian, and other races and ethnicities. Geographic distribution of mAb treatment was illustrated using zip code of patient residence.^21,22^

To evaluate comparative effectiveness, the primary outcome was hospital-free days up to day 28 after mAb treatment. This outcome is an ordinal endpoint with death up to day 28 as the worst outcome (labeled -1), then the length of time alive and free of hospital, such that the best outcome would be 28 hospital-free days. If a patient had intervening days free of hospital and was then re-hospitalized, the patient was given credit for the intervening days as free of the hospital. Secondary outcomes included mortality at 28 days. We evaluated rates of hospitalization by infusion location, and incidence of adverse events (**Supplement**). We assessed SARS-CoV-2 variant prevalence in our Pennsylvania catchment over time using Global Initiative on Sharing All Influenza Data (GISAID) data.^23^

### Data Collection

OPTIMISE-C19 was embedded in the EHR to access routine patient care data and was augmented by manual review and data collection. We ascertained the primary outcome of hospital-free days by linking inpatient (Cerner, Kansas City, Missouri) and outpatient (Epic, Madison, Wisconsin) EHRs, as shown in prior work.^18^ We conducted patient-directed phone calls at day 28 to ascertain health care encounters outside our health system, and Social Security Administration Death Master File queries for vital status.^24^ We collected adverse events in i.) a secure, electronic file sharing application completed by infusion center nurses on the day of mAb treatment, and ii.) an internal patient safety reporting system for adverse reactions completed by nursing and physician staff in infusion centers and EDs. Adverse event severity was adjudicated blinded to mAb type.

### Statistical Analysis

To determine the epidemiology and equitability of mAb infusions in our region, we measured the proportion of EHR-screen eligible patients treated with any mAb stratified by demographics, geography, and prior to or after trial launch. Treatment “prior to the trial” was from December 9, 2020 until March 9, 2021, and “during the trial” from March 10 to June 25, 2021.

To analyze comparative effectiveness, the trial statistical analysis plan was written by blinded investigators prior to data lock and analysis (**Supplement**) and applied to patients treated with mAb from March 10, 2021 to June 25, 2021. The platform is designed to continuously evaluate multiple mAb, with randomization continuing until pre-determined statistical thresholds are met. The trial launched with equal allocation randomization and planned interim analyses for adaptive randomization where mAb performing better would be given higher randomization probabilities. The mAb arm at the first adaptive analysis with the largest sample size was specified as the referent arm, as there was no non-mAb control and all patients received active treatment. All mAb were compared to each other. The methods and results are reported in accordance with the CONSORT Pragmatic Extension checklist (**Supplement**).^25^ An unblinded statistical analysis committee conducted interim and final analyses with R version 4.0.5 using the RStan package version 2.21.0 (R Foundation, Vienna, Austria) and reported results to the UPMC Chief Medical Officer who functioned in a data and safety monitor role for the study.

The primary analysis population was the “as-infused” population which consisted of patients randomly allocated mAb and subsequently treated. As all trial arms included mAb, there was no anticipated relationship between lack of infusion and the assigned arm. Baseline patient characteristics of those infused were compared to those allocated but not infused using mean (SD) or median (IQR) for continuous variables, and proportions for categorical variables.

The primary analysis model was a Bayesian cumulative logistic model that adjusted for treatment location (infusion center or ED), age (<30, 30-39, 40-49, 50-59, 60-69, 70-79, and ≥80 years), sex, and time (2-week epochs). The comparison between individual mAbs were based on the relative odds ratio between a given two arms for the ordinal primary outcome. An odds ratio for an arm to a comparator greater than 1 implies improved outcomes on the ordinal scale. A sliding scale of equivalence was set according to odds ratio bounds of 0.25, 0.20, 0.15, 0.10, and 0.05. Equivalence between two arms was defined as a 95% posterior probability that the odds ratio is within a given odds ratio bound. Inferiority of one arm compared to another was defined as a 99% posterior probability of an odds ratio less than 1.

To determine potential differential treatment effects of mAbs by Covid-19 variant type, patients were categorized into four time epochs relative to variant prevalence in 2021 (March 10 – 31; April 1 – April 30; May 1 – 31; June 1 – 25). Treatment effects were measured from the primary model over time with hierarchical prior distributions.

### Decision to publish interim results

The U.S. Department of Health and Human Services halted distribution of bamlanivimab alone on March 25, 2021, and of bamlanivimab-etesevimab on June 25, 2021, due to concern of lack of efficacy against certain SARS-CoV-2 variants that were predominant in the U.S. at those times.^26,27^ As further treatment with these mAbs was not possible, we unblinded and analyzed patients allocated through June 25, 2021, with follow up completed by July 23, 2021.

## RESULTS

### Expansion of monoclonal antibody treatment

Prior to the trial, 16,345 patients were EHR-screen eligible. Of these, 502 patients overall (3.1%) were treated with mAb, of whom the proportion of Black (2.6%) and White (3.1%) treated patients were similar. After trial launch, 1,201 of 5,173 EHR-screen eligible patients (23.2%) were treated. The proportion of eligible White patients receiving mAb increased from 3.1 to 21.6% and eligible Black patients receiving mAb increased from 2.6 to 29.9% during the trial. Broad geographic expansion was evident across the catchment (**Figure 1**).

**Figure 1.**
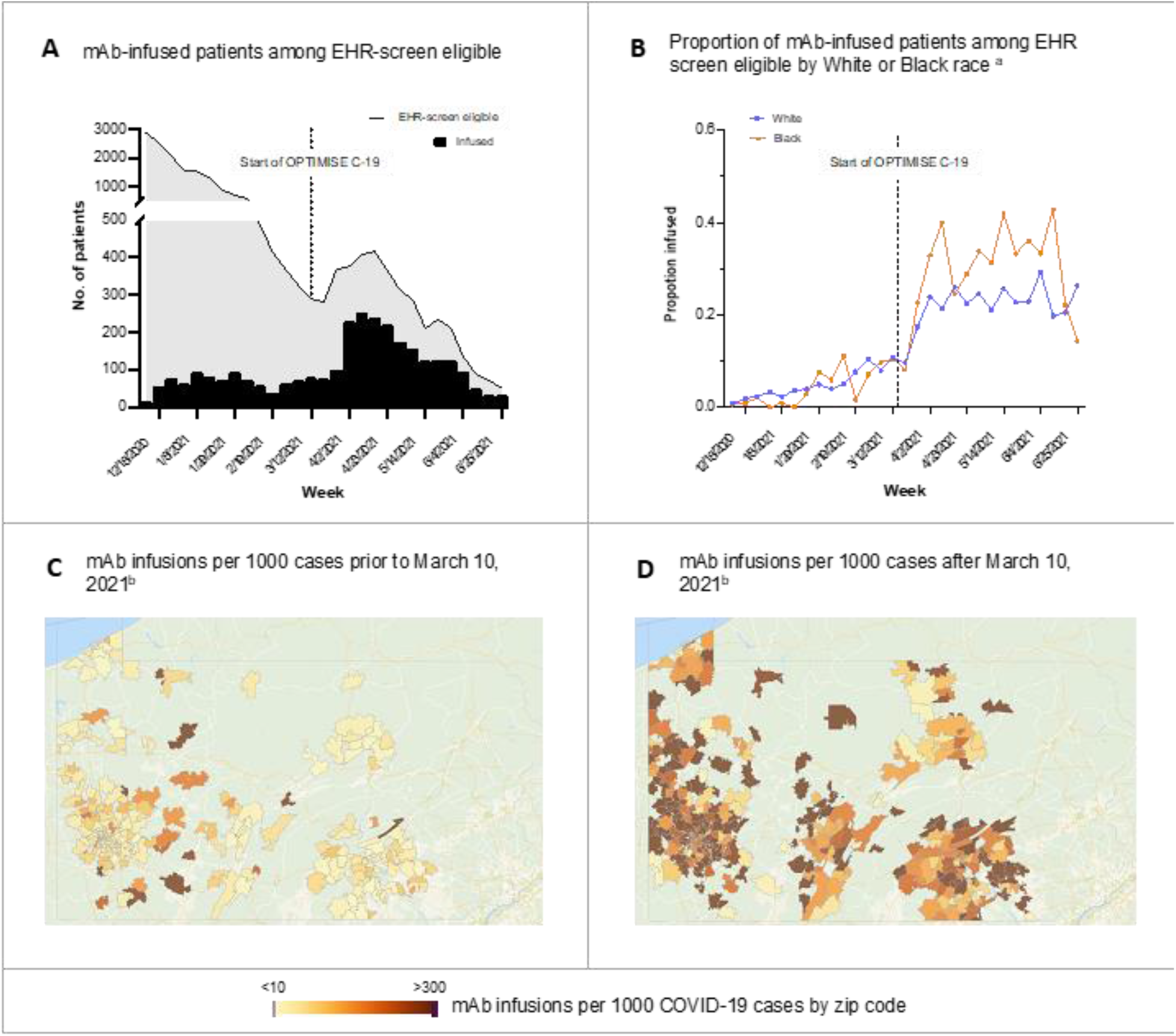
Epidemiology of Monoclonal Antibody Infusions. Panel A shows the number of mAb-infused patients among the EHR screen eligible. Panel B shows the proportion of mAb-infused patients among EHR screen eligible by White or Black race. Hispanic ethnicity, Other, and Unknown race are not shown due to small sample sizes. Panel C shows the number of mAb infusions per 1,000 cases prior to March 10, 2021. Panel D shows the number of mAb infusions per 1,000 cases after March 10, 2021. Zip codes with <10 Covid-19 cases or those outside of Pennsylvania are not shown. *Interpretive example:* Prior to the trial (*panel A*), mAb infusion was low. In panel B, the proportion of White and Black race infused among EHR-screen eligible patients increased. In the UPMC catchment in Pennsylvania, mAb infusions also increased in amount (darker, *panel C, D*) and in geographic distribution (more zip code areas colored) during OPTIMISE-C19.

### Trial patients

Between March 10 and June 25, 2021, 5,173 outpatients with a positive SARS-CoV-2 test within UPMC were EHR-screen eligible, of whom 1,382 were referred and underwent random mAb allocation. 1,084 EUA-eligible patients with a positive SARS-CoV-2 test from outside of UPMC were also referred, yielding a total of 2,466 patients who were assigned a random mAb allocation. Of these, 1,935 (78%) were infused and comprised the primary analysis cohort (bamlanivimab [n = 128], bamlanivimab-etesevimab [n = 885], casirivimab-imdevimab [n = 922], **Figure 2**). Of the 531 patients assigned a random mAb allocation and excluded from the primary analysis, most (n = 300, 56%) were not infused due to becoming clinically ineligible at time of infusion scheduling (n = 154, 29%) or patient declining treatment (n = 146, 27%). Eighty-eight patients (17%) received mAb at a location without available EHR data or while hospitalized for non-Covid-19 reasons. No patient or provider requested a specific mAb different than randomized assignment.

**Figure 2.**
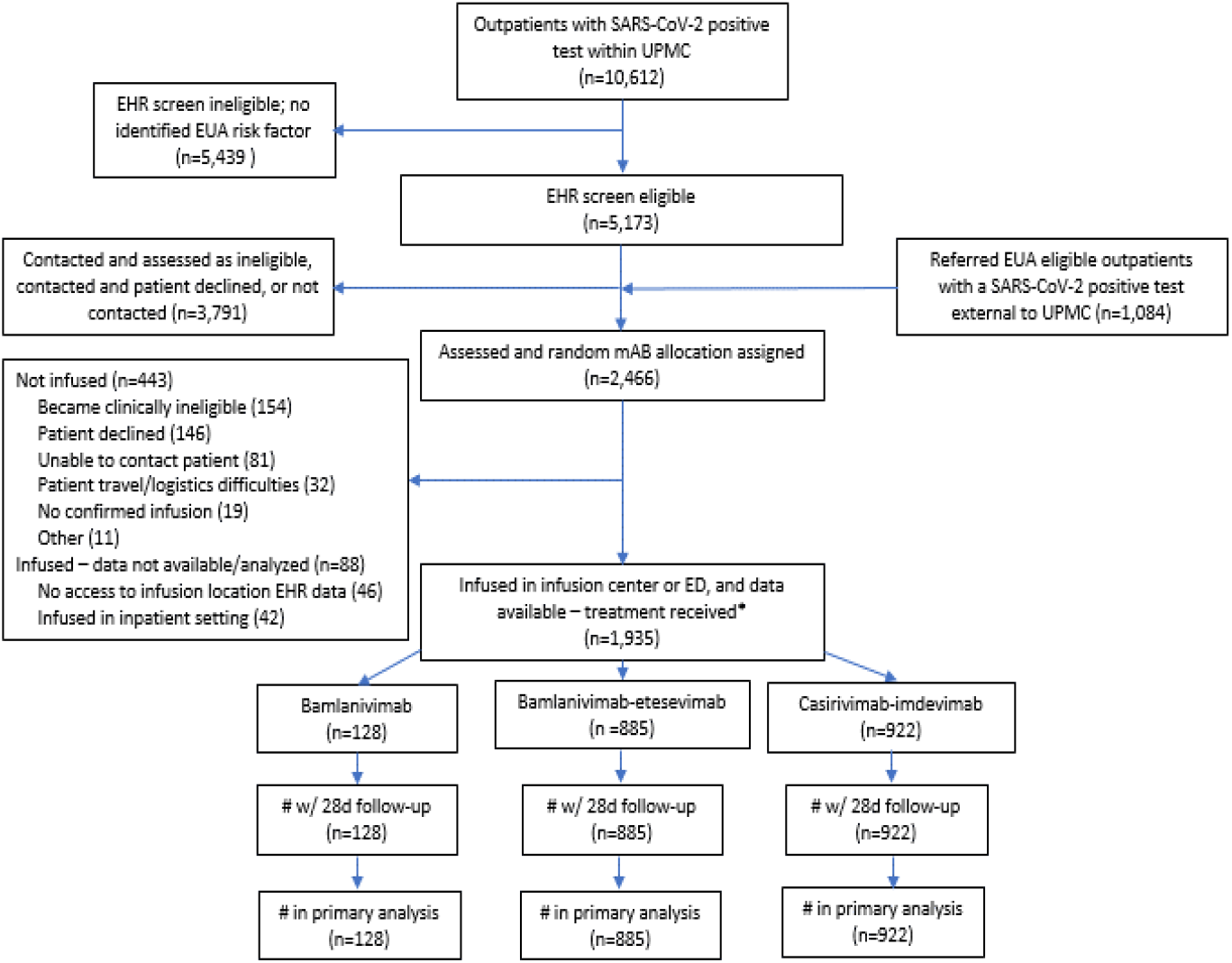
CONSORT Diagram. Due to pharmacy logistics, 5 patients who received bamlanivimab-etesevimab had been randomly assigned to casirivimab-imdevimab, and 7 patients who received casirivimab-imdevimab had been randomly assigned to bamlanivimab-etesevimab. All infused patients who received bamlanivimab monotherapy had been randomly assigned to bamlanivimab monotherapy. The FDA mAb policies changed over time, resulting in varying mAb availability and EUA eligibility criteria over time (**Supplement**).

Baseline characteristics were similar across groups (**Table 1**). The mean age for the three groups was 55 - 57 years, half were female (range, 53% - 54%), 18% were Black (range 12% - 19%), and the most common risk factors were advanced age, high body mass index, and hypertension. Of the 241 patients for whom vaccine status was known, 57 (24%) reported being unvaccinated, 120 (49%) partly vaccinated, and 64 (27%) fully vaccinated. Mean duration of symptom onset to referral was 5 (2.1) days.

**Table 1.**
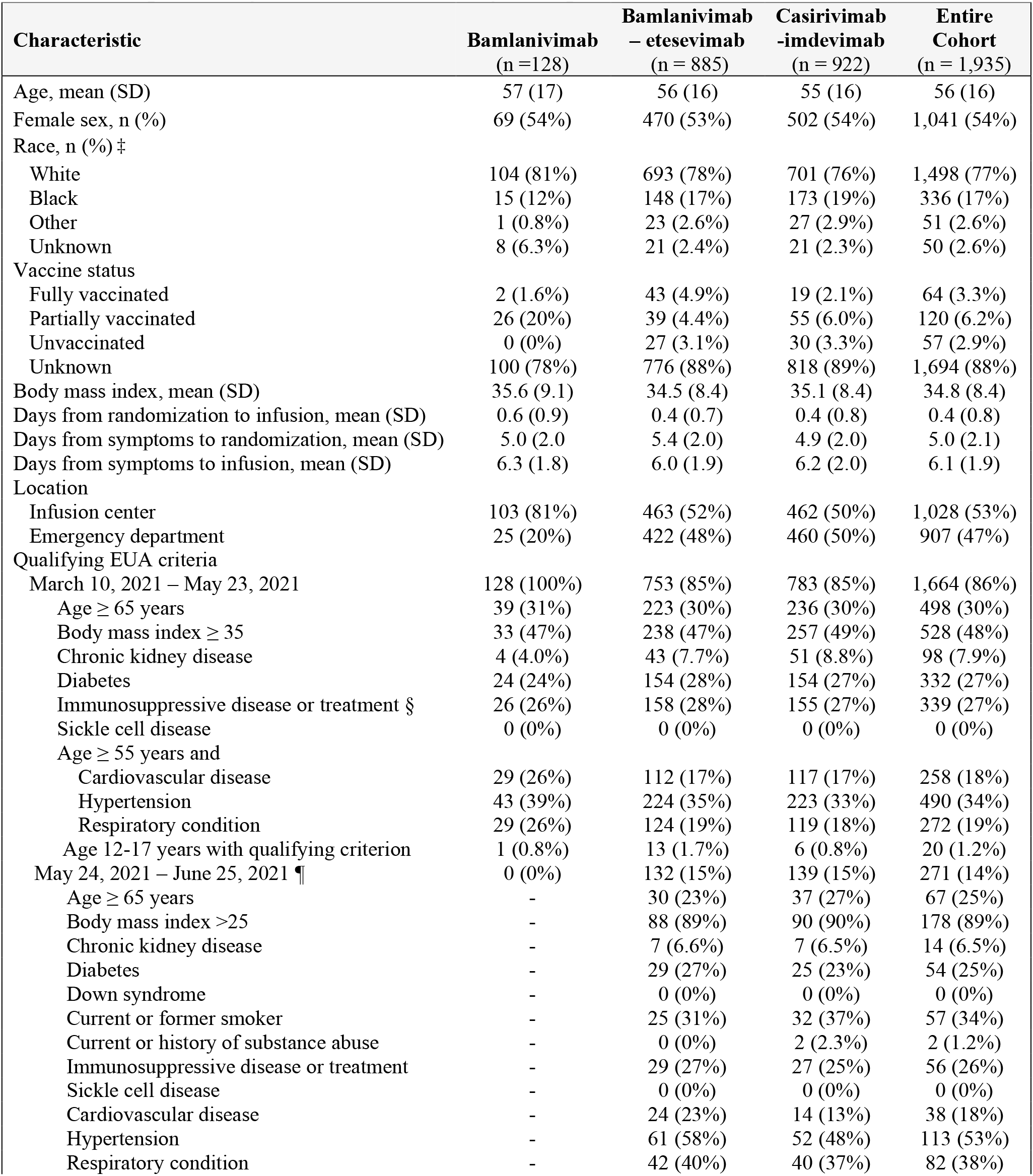
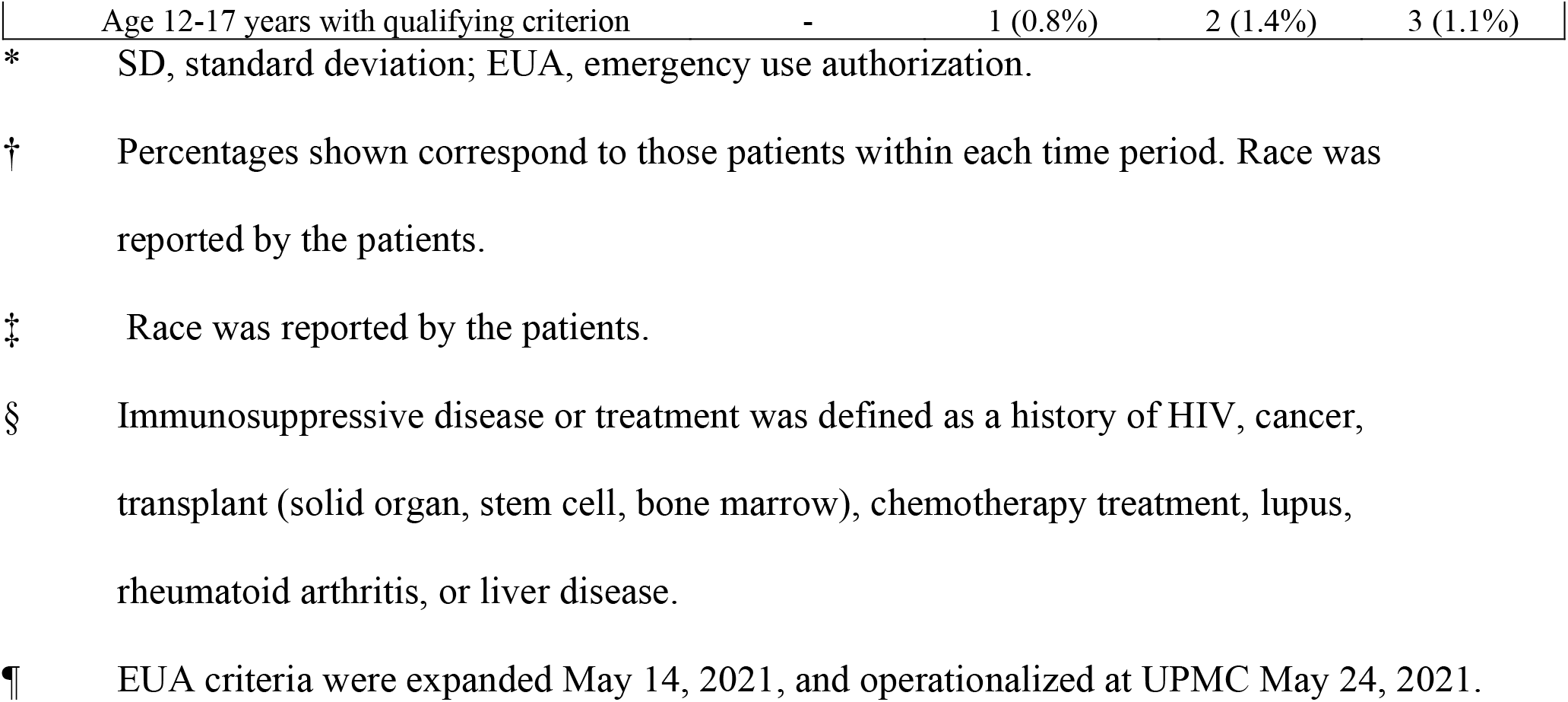
Demographic and Clinical Characteristics of the Patients at Baseline and Comparison by Monoclonal Antibody Group.

### Primary outcome

The median hospital-free days were 28 (IQR, 28-28) for each mAb group (**Table 2, Figure 3)**. Relative to the casirivimab-imdevimab group, the posterior median adjusted odds ratios from the primary model were 0.58 (95% credible interval, 0.30 to 1.16) and 0.94 (95% credible interval, 0.72 to 1.24) for the bamlanivimab and bamlanivimab-etesevimab groups, respectively. The probabilities of inferiority for bamlanivimab versus bamlanivimab-etesevimab and casirivimab-imdevimab were 91% and 94% respectively. The probability of equivalence between bamlanivimab-etesevimab and casirivimab-imdevimab with a bound of 0.25 for the odds ratio was 86%. No comparison met prespecified criteria for statistical inferiority or equivalence.

**Table 2.**
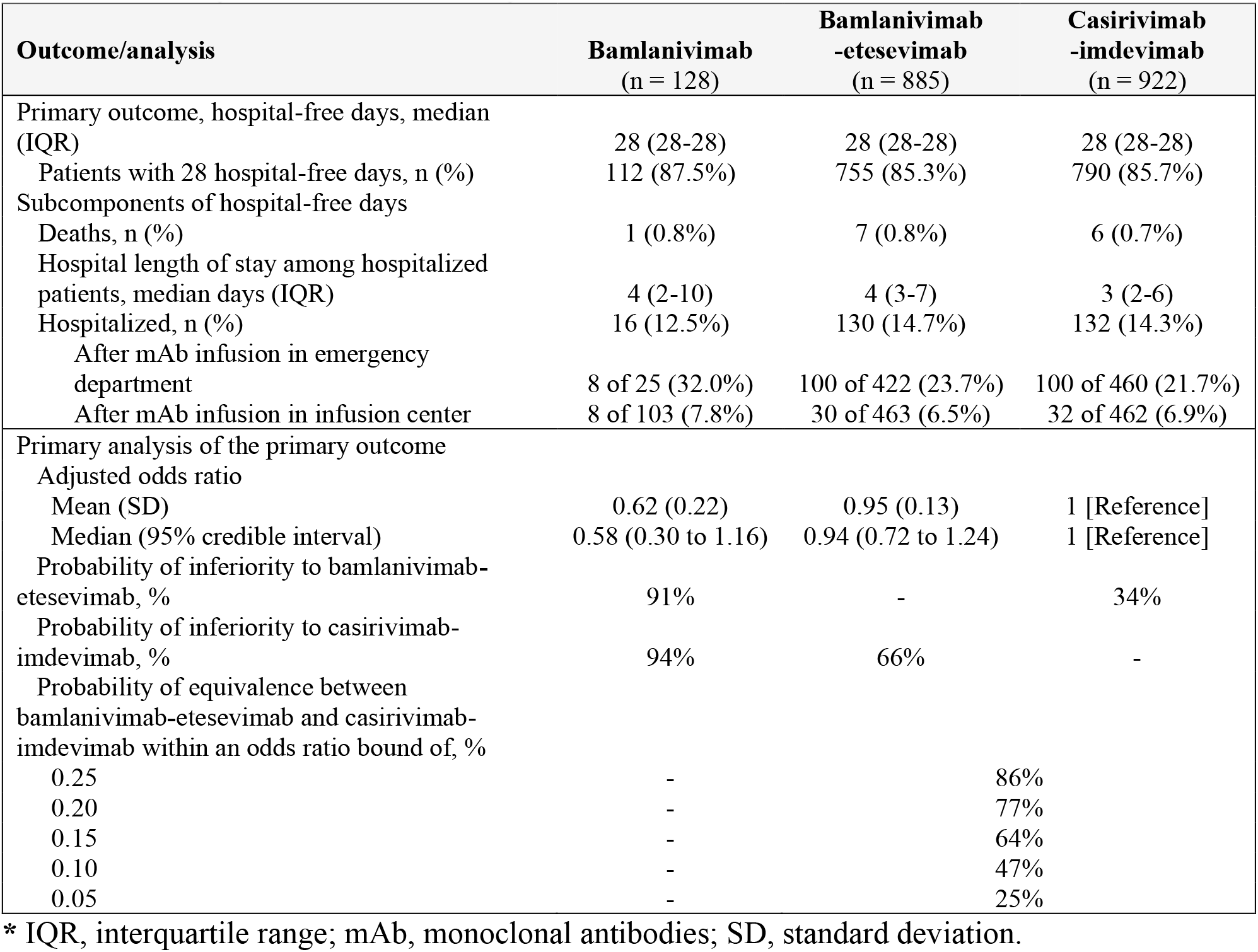
Primary Outcomes and Analysis.

**Figure 3.**
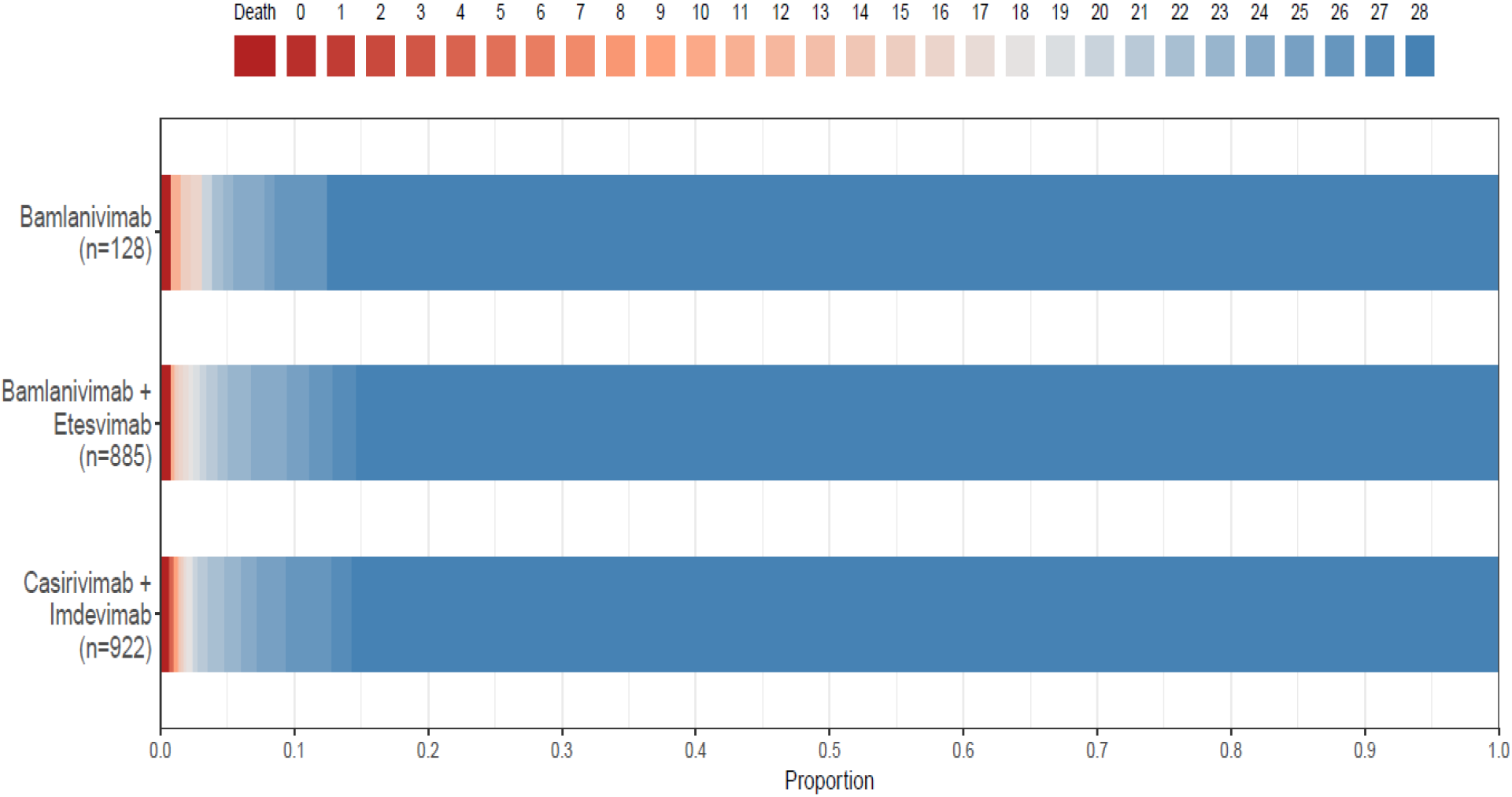
Hospital-Free Days to Day 28. Primary outcome is displayed as horizontally stacked proportions by monoclonal antibody type. Red represents worse values and blue represents better values. The median adjusted odds ratio from the primary analysis, using a Bayesian cumulative logistic model, were 0.58 (95% credible interval, 0.30 to 1.16) and 0.94 (95% credible interval, 0.72 to 1.24) for the bamlanivimab and bamlanivimab-etesevimab groups compared with the casirivimab-imdevimab group. These odds ratios yielded 91% and 94% probabilities of inferiority of bamlanivimab versus bamlanivimab-etesevimab and casirivimab-imdevimab respectively, and an 86% probability of equivalence between bamlanivimab-etesevimab and casirivimab-imdevimab at an odds ratio bound of 0.25.

### Secondary outcomes

The 28-day mortality rates were 0.8% (1/128), 0.8% (7/885), and 0.7% (6/922) and hospitalization rates were 12.5% (16/128), 14.7% (130/885), and 14.3% (132/922), in the bamlanivimab, bamlanivimab-etesevimab, and casirivimab-imdevimab groups, respectively (**Table 2**). For patients receiving mAb in an outpatient infusion center, rates of hospitalization after treatment were 7.8% (bamlanivimab), 6.5% (bamlanivimab-etesevimab), and 6.9% (casirivimab-imdevimab). For patients receiving mAb in an ED, rates of hospitalization after treatment were 32% (bamlanivimab), 23.7% (bamlanivimab-etesevimab), and 21.7% (casirivimab-imdevimab).

### Adverse events

Twenty-one infusion-related adverse events occurred in 0% (0/128), 1.4% (12/885), and 1.0% (9/922) of patients treated with bamlanivimab, bamlanivimab-etesevimab, and casirivimab-imdevimab, respectively. Five events (0 bamlanivimab, 1 bamlanivimab-etesevimab, 4 casirivimab-imdevimab) were adjudicated as serious (**Supplement**).

### Differences in treatment over time

During the trial the Alpha SARS-CoV-2 variant was the dominant variant of concern, while the Delta variant became more prevalent in the final time epoch (**Supplement**). We found no relative difference in mAb treatment effects over time and no comparisons reached a pre-specified statistical threshold (**Supplement**).

## DISCUSSION

In a learning healthcare system trial of mAb for Covid-19, treatment of EHR-screen eligible patients increased 7.5-fold, particularly among historically and geographically underserved populations. We found a 91% and 94% probability of inferiority of bamlanivimab respectively to bamlanivimab-etesevimab and casirivimab-imdevimab, and an 86% probability of equivalence between bamlanivimab-etesevimab and casirivimab-imdevimab at the first pre-specified bound, with regard to the odds of improvement in hospital-free days by 28 days. However, the bamlanivimab and bamlanivimab-etesevimab arms were stopped early, and the identified probabilities did not meet prespecified statistical triggers for trial conclusions of inferiority or equivalence.

OPTIMISE-C19 is designed to continuously compare all available mAb for Covid-19 and can stop mAb arms based on internal statistical triggers or external factors, such as U.S. federal decisions limiting mAb availability or re-introduction of mAb into distribution networks. Had federal decisions not prompted unblinding, the internal action would have been to generate updated randomization proportions and continue enrollment. The trial is currently evaluating casirivimab-imdevimab and sotrovimab.^28,29^

Systematic inequities have exacerbated racial health disparities throughout the Covid-19 pandemic, and a focused effort on equitable, expanded access to life-saving treatments is crucial for population health.^30^ This trial was able to increase access using a multifaceted approach: 1) infrastructure with infusion locations in all geographic regions, 2) comprehensive, electronic screening of all patients within the system with a positive SARS-CoV-2 followed by direct-to-patient outreach phone calls, 3) paper referral form for patients without access to an in-system provider, 4) community leader and existing outreach network collaboration for patient engagement, 5) phone line for patients and community members to call and speak with a healthcare professional, 6) providing infusions at home for patients without transportation.

The finding of potential inferiority of bamlanivimab is similar to mechanistic studies that suggest a waning efficacy of bamlanivimab in the face of certain SARS-CoV-2 variants. It supports the FDA decision to revoke the bamlanivimab EUA.^26^ A recent observational study reached a different conclusion and found similar effectiveness between bamlanivimab and casirivimab-imdevimab.^31^ However, this study analyzed patients treated between November 2020 and February 2021, prior to widespread emergence of variants and U.S. federal decisions to halt bamlanivimab distribution.

SARS-CoV-2 variant epidemiology changes rapidly and this report addresses a period when the Alpha variant was dominant, and the Delta variant was not widespread. Notably, the U.S. federal government reinstated circulation of bamlanivimab-etesevimab on August 27, 2021 in areas where variant resistance to this mAb is < 5%, based on *in vitro* data of activity against the Delta variant, and lack of activity against the Beta, Gamma, Delta plus, and B.1.621 variants.^31,32^ The similar effectiveness of bamlanivimab-etesevimab and casirivimab-imdevimab in the current trial supports this decision. Also aligning with these data is a recent observational study of 165 patients that found no difference in hospitalization or death between bamlanivimab-etesevimab and casirivimab-imdevimab in patients infected with Alpha, but worse outcomes with bamlanivimab-etesevimab in patients infected with Gamma.^33^

The strengths of this report include capture of nearly all patients infused with mAb from 49 sites across a large geographic region, enhancing the generalizability of the results. In addition, an advantage of the Bayesian design is that any data, including data following unplanned cessation in enrollment into a trial arm, can be analyzed and quantified as posterior probabilities, which is potentially more useful and is more quantitative than a frequentist conclusion of failure to reject a null hypothesis possibly because of lack of power.^34^ Third, the trial was embedded into usual care which enhanced patient and provider engagement.^12^ The trial also has limitations. First, the results are presented before any prespecified internal trigger was reached. Nonetheless, to our knowledge, this trial represents the largest randomized comparative effectiveness data of mAb for Covid-19. Second, the absence of patient-level variant data limited ability to directly assess comparative effectiveness relative to variant strains. Alpha was also the dominant variant during the majority of the trial. Using regional data as a surrogate for variant data in the Pennsylvania population over time, we found no difference in treatment effect over time. Third, we primarily relied on UPMC EHR data to capture death and hospitalization, and patients may have accessed care outside our health system after mAb treatment. We conducted direct-to patient calls and national death registry queries to address this concern. Fourth, the EHR eligibility screen identified most, but not all EUA risk factors, and could not identify if a patient was symptomatic.

## Conclusion

In non-hospitalized patients with mild to moderate Covid-19, bamlanivimab, compared to bamlanivimab-etesevimab and casirivimab-imdevimab, resulted in 91% and 94% probabilities of inferiority with regards to odds of improvement in hospital-free days within 28 days. There was an 86% probability of equivalence between bamlanivimab-etesevimab and casirivimab-imdevimab at an odds ratio bound of 0.25. However, the trial was unblinded early to these mAb due to federal distribution decisions, and no mAb met prespecified criteria for statistical inferiority or equivalence.

## Data Availability

Data will not be made available.

## Acknowledgements

The authors thank the clinical staff of the UPMC monoclonal antibody infusion centers as well as the support and administrative staff behind this effort, including but not limited to: Michelle Adam, Jodi Ayers, Ashley Beyerl, Trudy Bloomquist, Mikaela Bortot, Jonya Brooks, Sherry Casali, Jeana Colella, Jennifer Dueweke, Jesse Duff, Janice Dunsavage, Jessica Fesz, Kathleen Flinn, Daniel Gessel, Amy Helmuth, Erik Hernandez, Larry Hruska, Allison Hydzik, Le Ann Kaltenbaugh, LuAnn King, Jim Krosse, Sheila Kruman, Amy Lukanski, Hilary Maskiewicz, Debra Masser, Katelyn Mayak, Rebecca Medva, Theresa Murillo, Melanie Pierce, Teressa Polcha, Kevin Pruznak, Debra Rogers, Rozalyn Russell, Sarah Sakaluk, Heather Schaeffer, Robert Shulik, Libby Shumaker, Susan Spencer, Betsy Tedesco, Ken Trimmer, Jennifer Zabala, and their entire teams. Donald M. Yealy, MD provided data and safety monitoring for the trial.

## Supplementary Appendix

This appendix has been provided by the authors to give readers additional information about their work.

Supplement to: **A learning health system randomized trial of monoclonal antibodies for Covid-19**

## Section 1 – Supplementary Methods

### Trial Design

Several problems may be encountered when generating robust clinical evidence, including barriers to conducting clinical trials, the generalisability of data from populations that are too broad or too narrow, the issue of equipoise especially when comparing different types of existing care, and the delay in translating results into clinical practice. A Randomized Embedded Multifactorial Adaptive Platform (REMAP) provides a strategy to address many of these problems by gaining economies of scale from a common platform, which allows for broad enrollment but retaining the ability to examine for heterogeneity of treatment effects between defined subgroups. A REMAP focuses predominantly on the evaluation of treatment options for the disease of interest that are variations within the spectrum of standard care (although testing of novel or experimental therapies is not precluded) and does so by embedding the trial within routine healthcare delivery. In this regard, the REMAP seeks to replace random variation in treatment with randomized variation in treatment allowing causal inference to be generated about the comparative effectiveness of different existing treatment options. The use of response adaptive randomization (RAR), which allows the allocation ratios to change over time based on accruing outcomes data, maximises the chance of good outcomes for trial participants. The embedding of such a platform within the day-to-day activities facilitates the translation of outcomes to clinical practice as a “self-learning” system. As such, it also functions as an embedded and automated continuous quality-improvement program. A final advantage of a REMAP for optimizing monoclonal antibody treatment outcomes is the ability to rapidly adapt to generate evidence if as new interventions emerge, avoiding the inevitable delays associated with conventional trials.

A REMAP applies novel and innovative trial adaptive design and statistical methods to evaluate a range of treatment options as efficiently as possible. The broad objective of a REMAP is, over time, to determine and continuously update the optimal set of treatments for the disease of interest. The set of treatments that may be tested within a REMAP comprise the set of all treatments that are used currently or may be developed in the future and used or considered for use in patients. The design maximizes the efficiency with which available sample size is applied to evaluate treatment options as rapidly as possible. A REMAP has the capacity to identify differential treatment effects in defined sub-groups (termed strata), address multiple questions simultaneously, and can evaluate interactions among selected treatment options. Throughout the platform, patients who are enrolled in the trial are treated as effectively as possible.

A conventional RCT (i.e., a non-platform trial) makes a wide range of assumptions at the time of design. These assumptions include the plausible size of the treatment effect, the incidence of the primary outcome, the planned sample size, the (typically, small number of) treatments to be tested, and that treatment effects are not influenced by concomitant treatment options. These assumptions are held constant until the trial completes recruitment and data are analyzed. Participants who are enrolled in a conventional RCT are not able to benefit from knowledge accrued by the trial because no results are made available until the trial completes. A REMAP uses five approaches to minimise the impact of assumptions on trial efficiency and maximises the benefit of participation for individuals who are enrolled in the trial.

These design features are:

- frequent adaptive analyses using Bayesian statistical methods
- response adaptive randomization
- evaluation of differential treatment effects in pre-specified sub-groups (strata)
- evaluation of specified intervention-intervention interactions
- testing of multiple interventions in parallel and, subsequently, in series

This creates a ‘perpetual trial’ with no pre-defined sample size, the objective of which is to define and continuously update best treatment over the lifetime of the REMAP. The design aspects, including the risk of type I and type II errors, are optimized prior to the commencement of the trial by the conduct of extensive pre-trial Monte Carlo simulations, modification of the trial design, and re-simulation in an iterative manner. The methods related to the application of the design features and the statistical analysis of this trial are outlined in the methods section of the protocol. A separate **Protocol Appendix** contains detailed versions of the protocol with relevant protocol amendments and the statistical analysis plan.

### Analytical Methods

Within the REMAP, two or more interventions are evaluated, and sequential Bayesian statistical analyses are used over time to incorporate new trial outcome information to determine if an intervention is superior, if one or more interventions are inferior, or if two or more interventions are equivalent, in comparison to all other interventions with respect to the primary endpoint. Every participant will be assigned an intervention. Inference in this REMAP is determined by analyses using pre-specified statistical models that incorporate time periods, age, and disease severity to adjust for heterogeneity of enrolled participants that might influence risk of death. These models incorporate variables that represent each intervention assigned to participants. The efficacy of each intervention is modeled as possibly varying in the different stratum in the REMAP. Whenever a model hits a predefined threshold for any of superiority, inferiority, or equivalence for an intervention with respect to the primary endpoint, this is termed a statistical trigger. At any given adaptive analysis, a statistical trigger may be reached for all participants or for one or more strata, further described in the statistical analysis plan (**Protocol Appendix, page 26**).

## Section 2 – Supplementary Tables

**Table S1.**
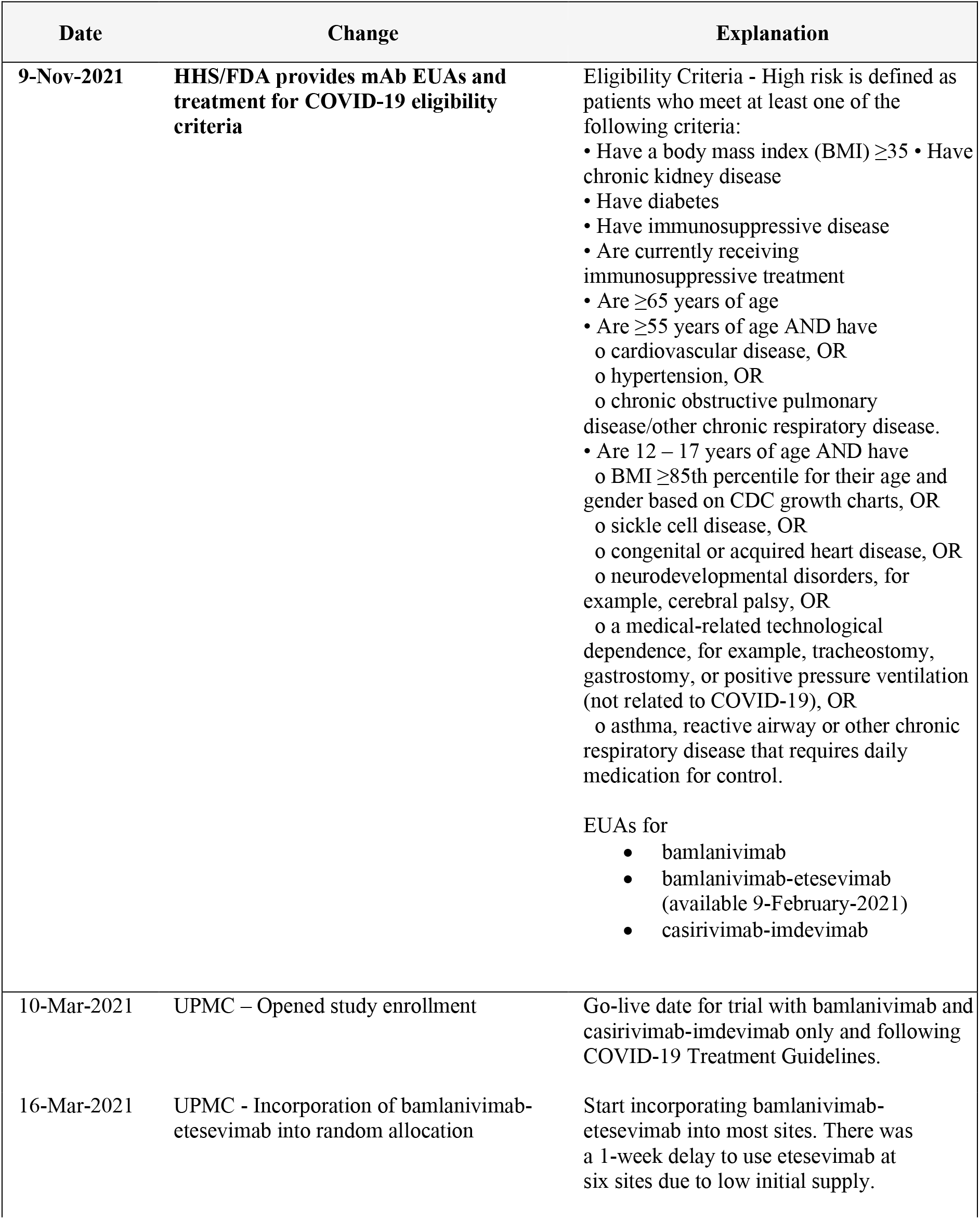
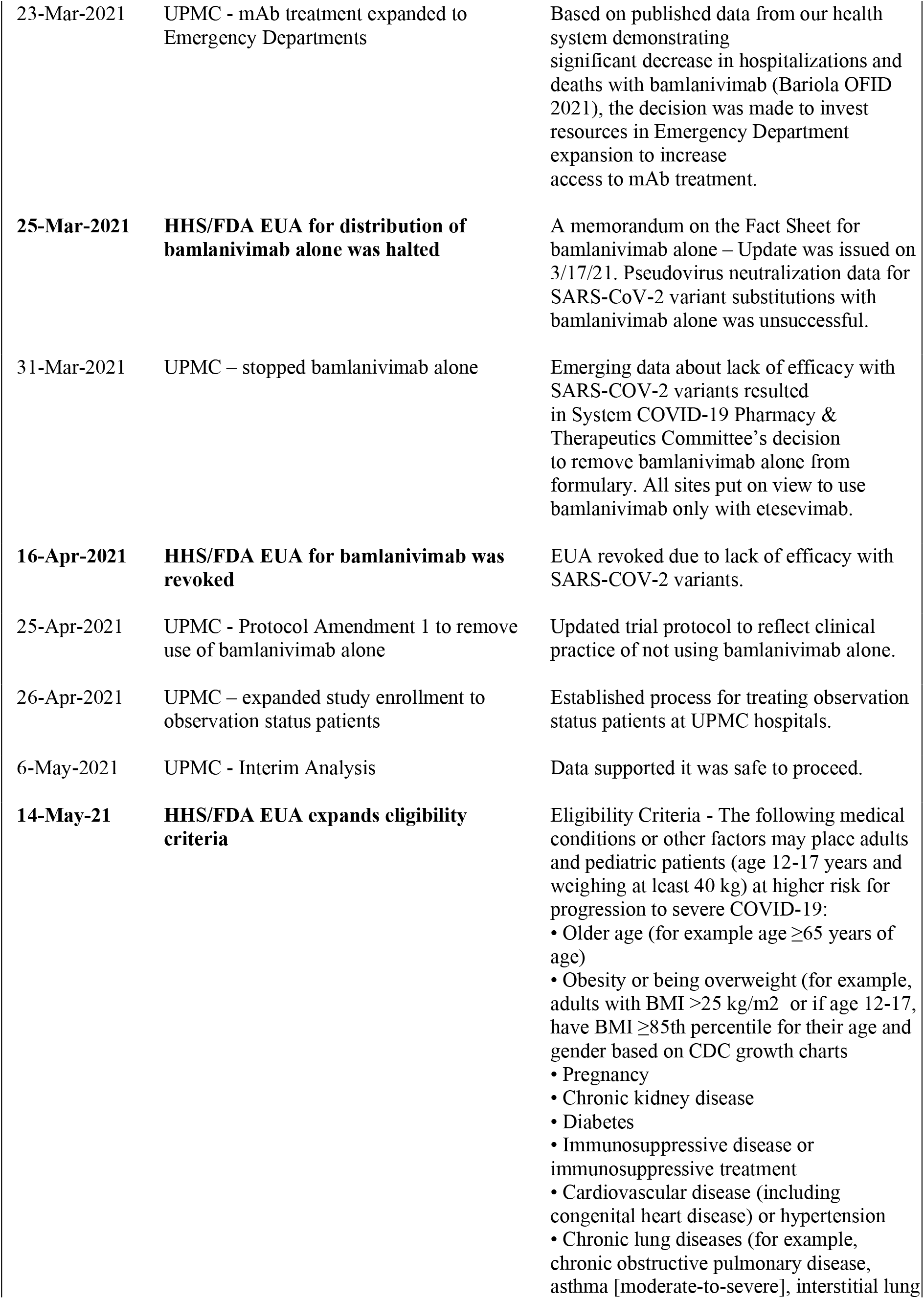
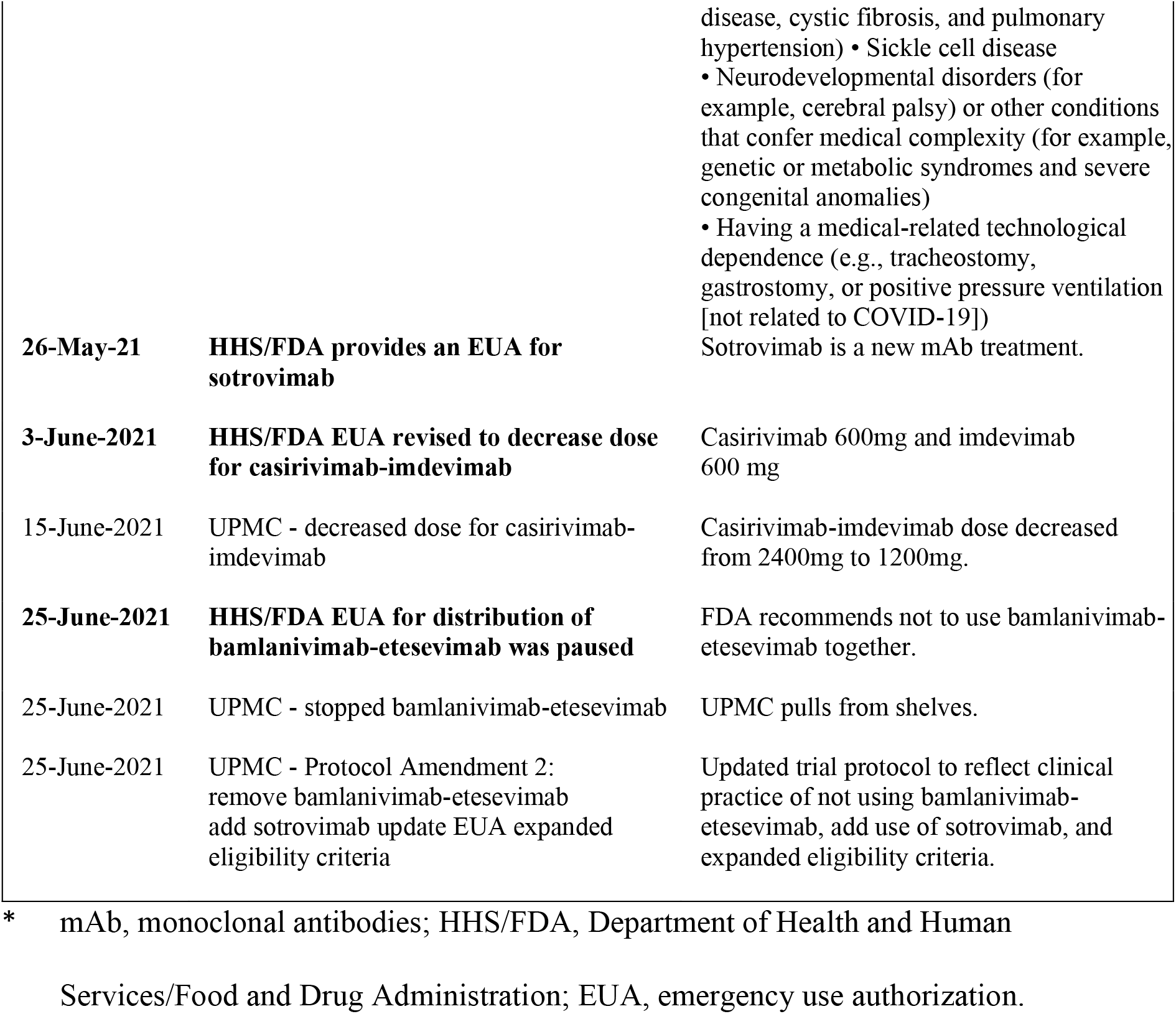
Department of Health and Human Services – Food and Drug Administration mAb Emergency Use Authorizations and UPMC Policy Changes Over Time.

**Table S2.** Adverse Events in Patients Receiving Monoclonal Antibody Treatment.

| Patient ID | mAb Received | Mild Reaction | Severe Reaction | Reaction Description |
| --- | --- | --- | --- | --- |
| 1 | Bamlanivimab + Etesevimab | Y |  | Hypoglycemic, hypotension: infusion stopped treated with diphenhydramine and fluid, patient discharged stable to home. |
| 2 | Bamlanivimab + Etesevimab | Y |  | Patient developed tightness in head/sinus area, and tightness in chest. Infusion not finished. Discharged home. |
| 3 | Bamlanivimab + Etesevimab | Y |  | Patient reported chest tightness, became lightheaded and dizzy. Infusion not finished. Discharged home. |
| 4 | Bamlanivimab + Etesevimab | Y |  | Patient developed hypotension; responded to 500mL LR, discharged home normotensive. |
| 5 | Bamlanivimab + Etesevimab | Y |  | Headache, responded to acetaminophen. |
| 6 | Bamlanivimab + Etesevimab | Y |  | Flushed, chest pain. Stopped infusion. |
| 7 | Bamlanivimab + Etesevimab | Y |  | Low back pain, radiating. Decreased infusion rate in half. Patient stated pain alleviated and denied any complaints |
| 8 | Bamlanivimab + Etesevimab | Y |  | Patient reported nausea, hot flash, severe lower back pain 1 minute into infusion. |
| 9 | Bamlanivimab + Etesevimab |  | Y | Patient stated flushed/throat "closing". Stopped infusion. Sent to ED for evaluation. |
| 10 | Bamlanivimab + Etesevimab | Y |  | Flushing and shortness of breath. |
| 11 | Bamlanivimab + Etesevimab | Y |  | Mild itchiness and redness at infusion site. |
| 12 | Bamlanivimab + Etesevimab | Y |  | Became itchy, developed hives, received diphenhydramine. |
| 13 | Casirivimab + Imdevimab | Y |  | Headache. |
| 14 | Casirivimab + Imdevimab |  | Y | Sudden onset nausea, vomiting, and weakness after 1 hour observation period; checked into ED and admitted post infusion |
| 15 | Casirivimab + Imdevimab | Y |  | Patient developed dizziness and chills which resolved prior to discharge. |
| 16 | Casirivimab + Imdevimab | Y |  | Right before discharge patient mentioned dizziness and lower blood pressure. Discharged to home. |
| 17 | Casirivimab + Imdevimab |  | Y | Hypertension and headache post-infusion, transferred to ED. |
| 18 | Casirivimab + Imdevimab |  | Y | Patient became hot, red face, reported chest pain 9/10. Oxygen saturation went from 95 to 85 to 73. Sent to ED. |
| 19 | Casirivimab + Imdevimab | Y |  | Chills, flushing, chest tightness, headache. Infusion stopped. |
| 20 | Casirivimab + Imdevimab |  | Y | Chest pain, shortness of breath, and back pain 2 minutes into infusion. Paramedics called. |
| 21 | Casirivimab + Imdevimab | Y |  | Shortness of breath, chest tightness 3 minutes into infusion. Medication stopped and symptoms resolved. |

## Section 4 – Supplementary Figures

**Figure S1.**
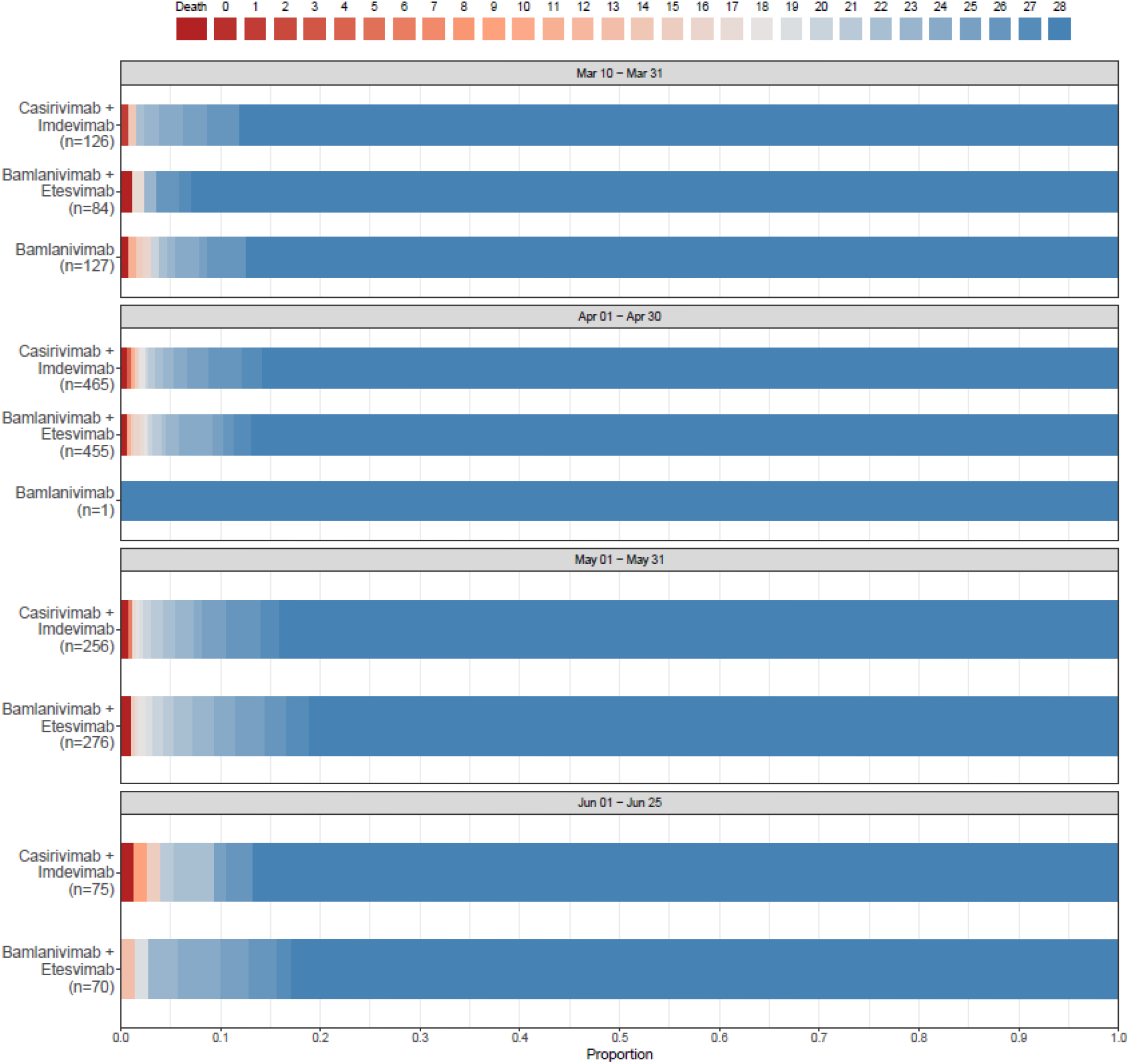
Treatment Heterogeneity Across Variant Date Prevalence Epochs.

**Figure S2.**
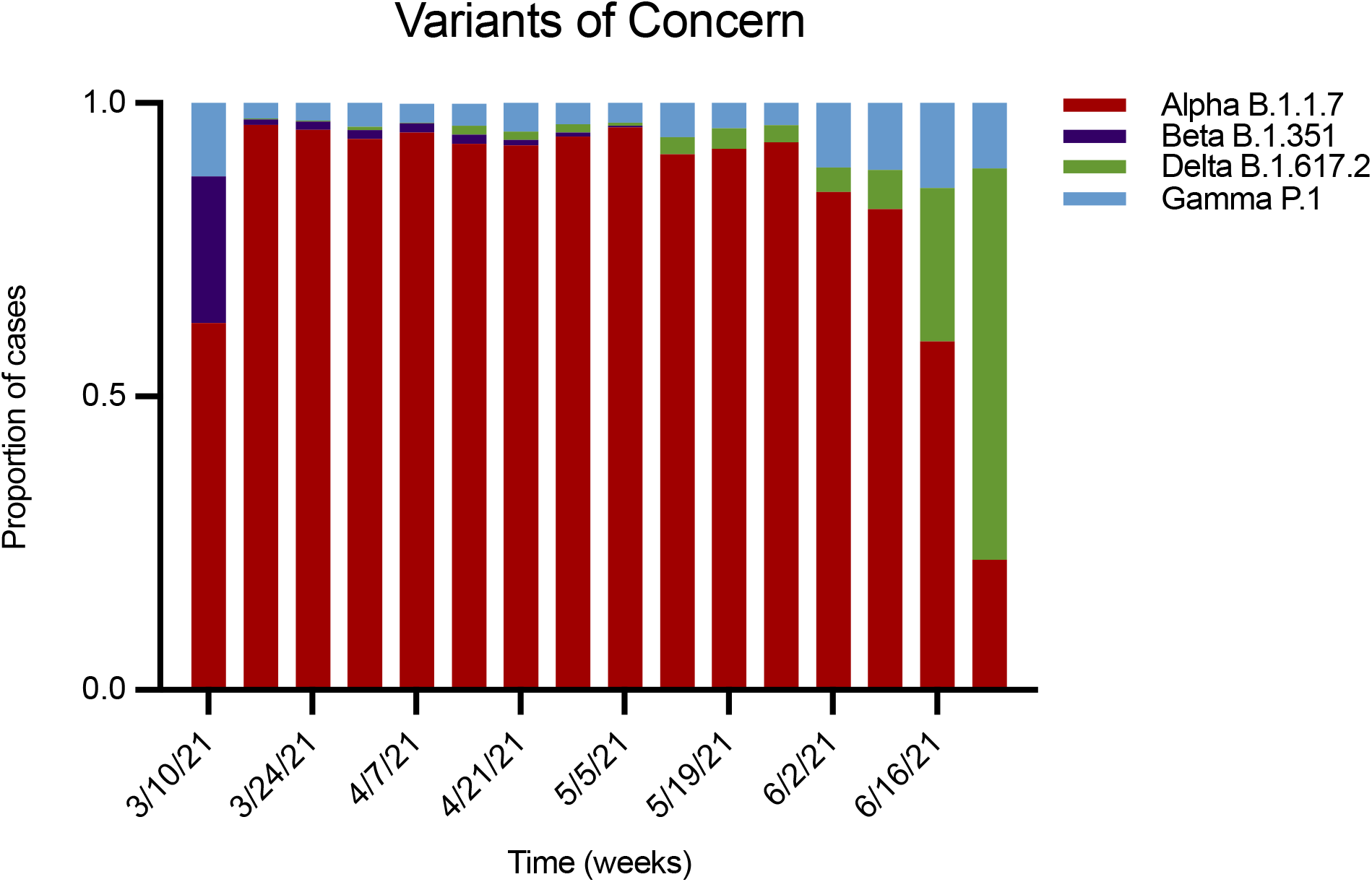
SARS-CoV-2 Variants of Concern Proportion in Pennsylvania During the Study.

**Figure S3.**
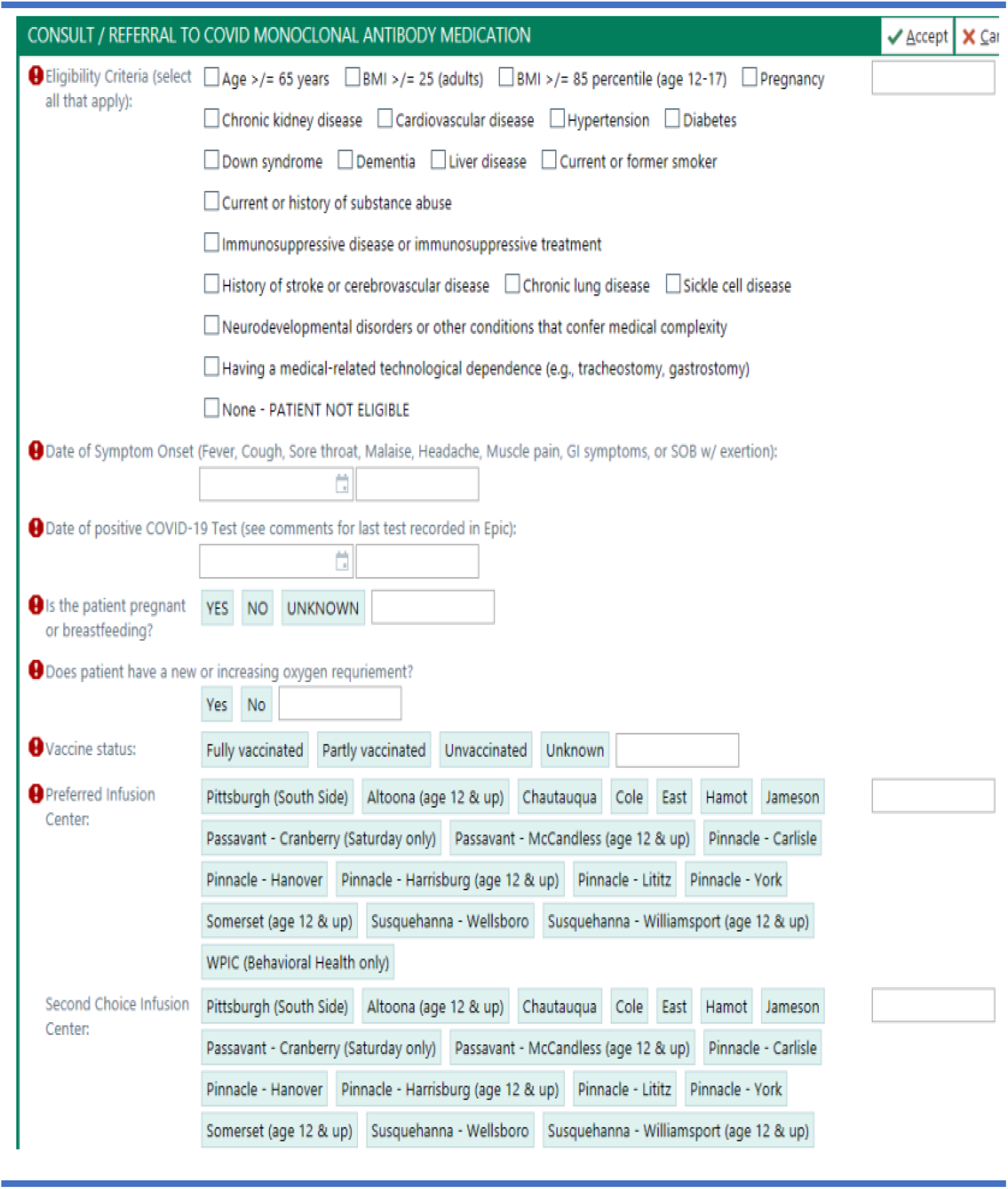
Monoclonal Antibody Referral Order in EPIC.

**Figure S4.**
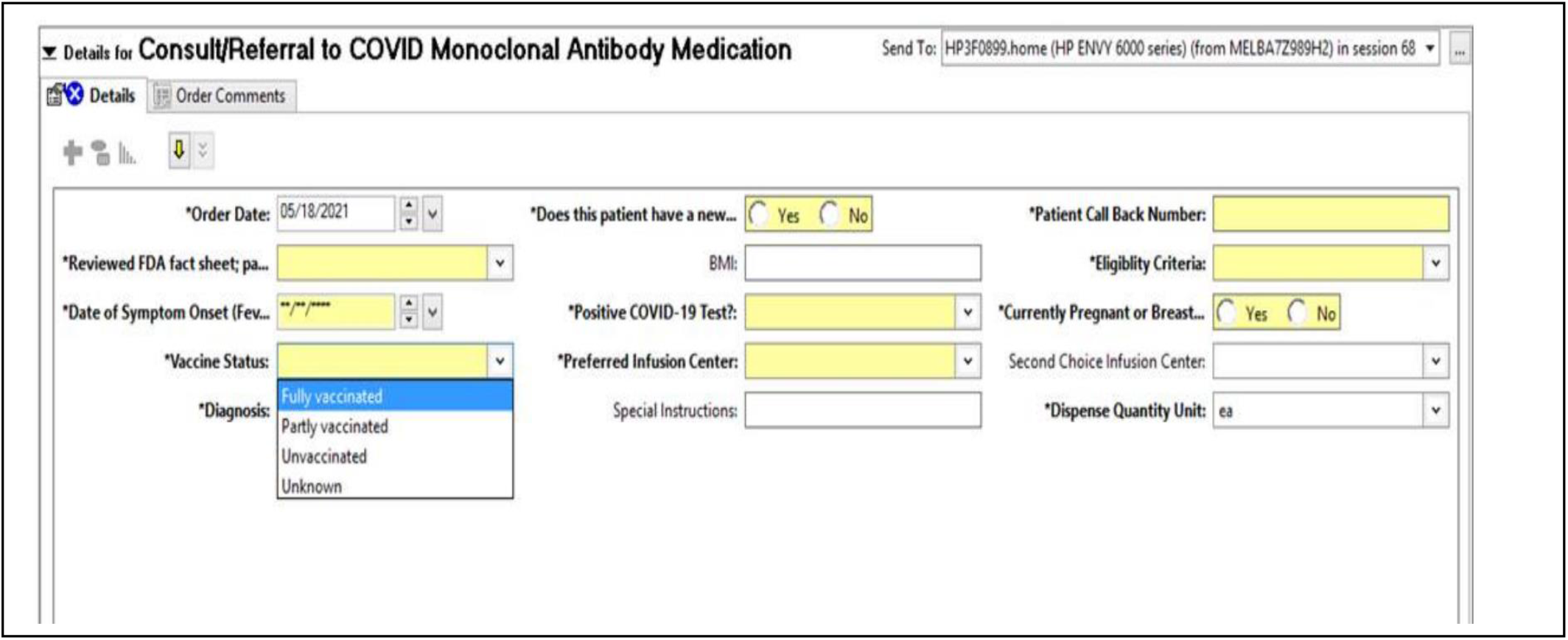
Monoclonal Antibody Referral Order in Cerner.

## Protocol Appendix

This trial protocol has been provided by the authors to give readers additional information about their work.

Protocol for: **A learning health system randomized trial of monoclonal antibodies for Covid-19**

## Protocol and Statistical Analysis Plan

This appendix contains the following items:

1. The original protocol document, final protocol document, and summary of changes.
2. The statistical analysis plan.

## ORIGINAL VERSION 1.0, February 24, 2021

A pragmatic evaluation of monoclonal antibody treatments in participants with COVID-19 illness (ClinicalTrials.gov, NCT04790786)

### Summary

#### Background

- FDA Emergency Use Authorizations (EUA) exist for multiple monoclonal antibodies (mAB) to treat COVID-19; the EUAs stipulate eligibility criteria, patient-physician communication, and clinical monitoring.^1^
- UPMC provides mABs as routine care; physicians order a mAB infusion and pharmacies assign whichever mAB is available under a therapeutic interchange approach. If scarcity exists, a lottery system is used.
- Physicians review with patients the EUA Fact Sheet for each mAB and explain they could be assigned any of the EUA-governed mABs.^2-4^

#### Approach

- Structure the therapeutic interchange policy and lottery system using a UPMC pharmacy embedded assignment system that allows a comparative effectiveness evaluation of the multiple mABs.^5^
- Collect data from clinically performed UPMC processes and EUA requirements for routine care.
- Monoclonal antibodies (mAB) for COVID-19

#### Treatments Inclusion Criteria

These criteria are as per the FDA EUAs for COVID-19 mABs as of February 24, 2021.^1^

- Adult and pediatric patients (12 years of age and older weighing at least 40 kg) with a positive SARS-CoV-2 antigen or PCR test and within 10 days of symptom onset, and high risk of disease progression
- High risk is defined as patients who meet at least one of the following criteria:

- Body Mass Index (BMI) ≥35
- Have Chronic Kidney Disease
- Have diabetes
- Have immunosuppressive disease
- Are currently receiving immunosuppressive treatment
- Are ≥65 years of age
- Are ≥55 years of age AND have

- cardiovascular disease, OR
- hypertension, OR
- chronic obstructive pulmonary disease/other chronic respiratory disease
- Are 12-17 years of age AND have:

- BMI ≥ 85th percentile for their age and gender based on CDC growth charts, OR
- sickle cell disease, OR
- congenital or acquired heart disease, OR
- neurodevelopmental disorders (e.g., cerebral palsy),
- OR

- a medical-related technological dependence, for example, tracheostomy or gastrostomy), OR
- asthma, reactive airway, or other respiratory disease that requires daily medication for control.

#### Exclusion Criteria

These criteria are as per the FDA EUAs for COVID-19 mABs as of February 24, 2021.^1^

- Are hospitalized for the treatment of COVID-19
- Require oxygen therapy for the treatment of COVID-19
- Require an increase in baseline oxygen flow rate due to COVID-19 in those on chronic oxygen therapy due to underlying non-COVID-19 related comorbidity
- Have a known hypersensitivity to any antibody ingredient

#### Primary evaluation metric

Total hospital free days at 28 days

## 1. ABBREVIATIONS

AE: Adverse Events
B: bamlanivimab
B + E: bamlanivimab and etesevimab
BMI: Body Mass Index
C + I: casirivimab + imdevimab
CDC: Centers for Disease Control and Prevention
COPD: Chronic Obstructive Pulmonary Disease
COVID-19: Coronavirus disease 2019
CVD: Cardiovascular Disease
EUA: Emergency Use Authorization
FDA: Food and Drug Administration
HHS: Health and Human Services
HFD: Hospital Free Days
HTN: Hypertension
kDa: Kilodaltons
IgG1: Immunoglobulin G1
KG: Kilograms
mAB: Monoclonal Antibodies
PCR: Polymerase Chain Reaction
SAEs: Serious Adverse Events
SARS-CoV-Severe: Acute Respiratory Syndrome 2 Coronavirus 2
UATRC: UPMC Antibody Treatment and Evaluation Center

## 2. BACKGROUND and RATIONALE

## 3. BACKGROUND

While COVID-19 vaccination will reduce COVID-19-related morbidity and mortality, the learned immune response may vary between individuals. This means interventions such as monoclonal antibodies (mAB) will still be needed to prevent progression of COVID-19 illness. Monoclonal antibodies seek to mimic or enhance the natural immune system response against a pathogen and are often used in the care of patients with cancer or infection.

For viral infections, mABs are created by exposing a white blood cell to a particular viral protein, which is then cloned to mass produce antibodies to target that virus. For SARS-CoV-2, the virus that causes COVID-19, IgG1 mABs target the spike protein of SARS-CoV-2 and block viral attachment and entry into cells.

The SARS-CoV-2 mABs bamlanivimab and etesevimab, and the REGN-COV2 combination (casirivimab + imdevimab) reduce nasopharyngeal viral burden plus clinical outcomes including future emergency department visits and hospitalizations (Weinreich 33332778 NEJM, Gottlieb 33475701). Each received FDA Emergency Use Authorization (EUA) for use in selected populations (**Exhibit**). Additional trials of pre-exposure prophylaxis (NCT04497987) and other applications are underway, and additional mABs are in development.

The trials demonstrated the greatest impact of the REGN-COV2 dual therapy among patients who lacked neutralizing antibodies against SARS-CoV-2 at baseline and in those with high nasopharyngeal viral loads. Additionally, few patients in the bamlanivimab/etesevimab trial developed treatment-emergent SARS-CoV-2 resistance. This latter phenomenon may further enhance the need for therapies given the recent emergence of SARS-CoV-2 variants that may escape vaccination. However, the relative effectiveness of each mAB compared to the other is unknown, as is their effectiveness for emerging virus variants.

This Appendix to the UPMC Pilot Core (PittPro 20040210) describes the approach of the UPMC Antibody Treatment and Evaluation Center. We will conduct a pragmatic evaluation of monoclonal antibody treatments in participants with COVID-19 illness, starting with the patient population approved under the current FDA mAB EAUs.

## 4. RATIONALE

As of February 2021, there are over 60,000 new cases of COVID-19 diagnosed daily in the US https://covid.cdc.gov/covid-data-tracker/#trends_dailytrendscases, with over 7000 daily COVID-19 related hospital admissions Microsoft Power BI (powerbigov.us). Although case volumes are currently declining, COVID-19 remains a significant public health threat.

Despite the EUAs, the clinical use of mABs is low due in part to lack of patient access, complexities in drug allocation, and lack of knowledge among providers are contributing factors. Further, the comparative effectiveness of different mABs is unknown and not yet directly studied. The National Academies of Sciences, Engineering, and Medicine recently called for expanded access and clinical use of mABs, noting it is “critical to collect data and evaluate whether they are working as predicted”.

This evaluation seeks to expand access to mABs at UPMC and determine their relative effects versus each other, starting with those governed by EUAs.

## 5. OBJECTIVES AND METRICS

## 6. OBJECTIVES

The primary objective is to evaluate the clinical and biological effect of multiple monoclonal antibodies (mABs) in patients with COVID-19.

The primary hypothesis is clinical and biological effect will vary between mABs, by SARS-CoV-2 variants, and patient characteristics.

## 7. METRICS

The primary evaluation metric is total hospital free days (HFD) at 28 days after mAB receipt calculated as 28 minus the number of days during the index stay minus the number of days readmitted during the 28 days after treatment. Death within 28 days is recorded as -1 HFD. Secondary evaluation metrics include:

- All-cause and all-location mortality at 28 and 90 days
- Emergency department visits at 28 days
- Organ-support free days at day 28
- Where feasible:
  - SARS-CoV-2 nasopharyngeal and plasma viral loads among participants from baseline and longitudinally through day 28
  - SARS-CoV-2 antibody titers, antibody neutralization, and other immune responses at baseline and longitudinally through day 28
  - Detection of SARS-CoV-2 variants through next-generation sequencing at baseline and longitudinally through day 28
  - Determining the duration of SAR-CoV-2 infectivity and non-culture surrogates for SARS-CoV-2 infectivity among patients with persistent nasopharyngeal swab viral shedding

## 8. DESIGN

We will conduct a pragmatic evaluation of participants with COVID-19 illness under existing UPMC processes for the clinical care of COVID-19 positive patients, including EUA requirements for mAB administration. A patient who presents to a UPMC facility and tests positive for COVID-19 will, as per current common care, be offered monoclonal antibodies. Data that are already collected according to UPMC procedures and EUA requirements are used for analysis.

## 9. POPULATION

We will evaluate patients that present to UPMC Emergency Departments, urgent care sites, infusions centers and other facilities that can or do provide mABs for COVID-19. As of February 24, 2021, there are 3 EUAs, with common inclusion and exclusion criteria, and we will evaluate patients that meet these criteria. As other antibodies become available, we will modify this evaluation submission.

## 10. INCLUSION CRITERIA

As per the current EUA criteria, the following patients are included:

- Adult (> 18 years old)
- Children > 12 years old weighing at least 40 kg
- With a positive SARS-CoV-2 antigen or PCR test and within 10 days of symptom onset
- High risk of disease progression

High risk is defined as patients who meet at least one of the following criteria:

- A Body Mass Index (BMI) ≥ 35
- Have chronic kidney disease
- Have diabetes
- Have immunosuppressive disease
- Are currently receiving immunosuppressive treatment
- Are ≥ 65 years old
- Are ≥ 55 years of age AND have:
  - cardiovascular disease, OR
  - hypertension, OR
  - chronic obstructive pulmonary disease/other chronic respiratory disease
- Are 12-17 years of age AND have:
  - BMI ≥ 85^th^ percentile for their age and gender based on CDC growth charts, OR
  - sickle cell disease, OR
  - congenital or acquired heart disease, OR
  - neurodevelopmental disorders (e.g., cerebral palsy), OR
  - a medical-related technological dependence, for example, tracheostomy or gastrostomy), OR
  - asthma, reactive airway, or other respiratory disease that requires daily medication for control

## 11. EXCLUSION CRITERIA

As per the current EUA criteria, the following are excluded:

## 12. EVALUATED TREATMENTS

Patients will receive COVID-19 mABs governed by FDA EUAs, when their treating physician orders a mAB and they meet EUA criteria. Currently and under our examination, the treating physician do not choose a specific mAB product.

As of February 24, 2021, there are three such mABs as listed below.

## 13. BAMLANIVIMAB

Bamlanivimab is a human immunoglobulin G-1 (IgG1 variant) monoclonal antibody consisting of 2 identical light chain polypeptides composed of 214 amino acids each and 2 identical heavy chain polypeptides composed of 455 amino acids produced by a Chinese Hamster Ovary (CHO) cell line and molecular weight of 146 kDa.

## 14. BAMLANIVIMAB and ETESVIMAB

Etesevimab is a human IgG1 variant monoclonal antibody (mAb) consisting of 2 identical light chain polypeptides composed of 216 amino acids each and 2 identical heavy chain polypeptides composed of 449 amino acids produced by a Chinese Hamster Ovary (CHO) cell line and molecular weight of 145 kDa.

## 15. CASIRIVIMAB and IMDEVIMAB

Casirivimab, a human immunoglobulin G-1 (IgG1) monoclonal antibody (mAb), is covalent eterotetramer consisting of 2 heavy chains and 2 light chains produced by recombinant DNA technology in Chinese hamster ovary (CHO) cell suspension culture and has an approximate molecular weight of 145.23 kDa.

Imdevimab, a human IgG1 mAb, is a covalent heterotetramer consisting of 2 heavy chains and 2 light chains produced by recombinant DNA technology in Chinese hamster ovary (CHO) cell suspension culture and has an approximate molecular weight of 144.14 kDa.

## 16. CONCOMITANT THERAPY

All care and concomitant therapy are as per the treating providers.

## 17. CONDUCT

## 18. DATA COLLECTION

The EUAs require that healthcare facilities and providers report therapeutic information and utilization data through HHS Protect, Teletracking, or National Healthcare Safety Network as directed by the US Department of Health and Human Services.

We will collect data including baseline demographics and underlying conditions, results of SARS-COV-2 PCR or antibody testing, and initial care including mAB infusion completion. We will collect post-randomization healthcare encounters, including hospitalization, emergency department visits, ICU care, and other measures of healthcare utilization. We will use an electronic health record data collection process to augment existing UPMC data collection processes as necessary.

All data will be handled and secured as per University of Pittsburgh and UPMC data guidelines. There will be no research activities involving direct interaction with subjects performed as part of this evaluation.

In addition to the primary and secondary outcome data referenced in this submission, data collected will include the below areas. All data will be abstracted directly from the electronic health record and handled anonymously.

- Which mAB was administered, including date, time, and infusion completion as well as the location of the infusion
- Demographics (including age, sex, race, body weight, vaccination status)
- Healthcare encounters, including hospital and ICU admission status if applicable
- Medication usage and doses
- Hospital and ICU admission status, if applicable
- Administration of medications related to COVID-19, if applicable
- Remnant blood availability
- Laboratory and microbiology data, including COVID-19 testing done for clinical purposes

## 19. BIOSPECIMENS

Where feasible, we will collect discarded remnant blood samples and nasal/oropharyngeal swab samples to quantify the viral load and host response to the virus. As noted under data collection, we will record laboratory and microbiology data performed for clinical purposes.

## 20. ANTIBODY ADMINISTRATION

Antibodies will be administered as per the EUAs and UPMC Pharmacy and Therapeutics policies. Providers will explain mAB risks and benefits and provide the EUA Fact Sheets for Patients, Parents and Caregivers as per EUA requirements.

## 21. mAB assignment

The COVID-19 mABs are currently routinely used at UPMC. Once any order for mAB infusion is approved by the UPMC system oversight group, the pharmacy provides whichever EUA-governed mAB is available under a therapeutic interchange approach. Ordering physicians review with the patient the EUA Fact Sheet for each mAB and explain that the patient could receive any of the mABs governed by FDA EUAs.

If demand for mAB exceeds supply, UPMC has a lottery system to allot who receives the therapy once requested by a physician.

Our current proposal is a UPMC system quality improvement initiative, embracing and extending the current lottery system and therapeutic interchange policy for EUA-governed mABs for COVID-19 as follows:

1. The Physician orders mAB.
  a. If scarcity present and lottery system allow provision, proceed.
2. The Pharmacy fills order with one of the EUA-governed COVID-19 mABs using an embedded assignment system akin to current mAb provision. This system will allow a comparative effectiveness evaluation of the multiple mABs by effectively ensuring random allocation.
3. The Physician can agree to the assigned mAB or can request a specific mAB. It is the treating physician’s choice to accept the assigned mAB or not, and therefore patient consent for the mAB assignment is not required. Patients will be told which mAB they are receiving, along with an EUA Fact Sheet, as per EUA requirements.

## 22. STATISTICAL CONSIDERATIONS

## 23. STRATA

Predefined strata will include patients discharged home after infusion, patients admitted to hospital after infusion, prior vaccination, and if known, presence of virus variants of concern at baseline and presence of neutralizing antibodies to SARS-CoV-2 at baseline.

## 24. NUMBER of PARTICIPANTS

Sample size is determined by case volume throughout the course of the pandemic.

## 25. STATISTICAL ANALYSIS

The primary evaluation metric is the number of days free from hospitalization to day 28. We will finalize a statistical analysis plan which will consider mAB assignment, heterogeneity of treatment effect by patient characteristics and virus variants, and interaction with other treatments. Due to uncertainty in sample size, we will use a Bayesian adaptive design to ensure ability to provide statistical inference despite variable sample size.

## 26. ETHICAL CONSIDERATIONS

## 27. DATA MONITORING

Evaluation center leadership will regularly monitor monthly reports on enrollment, patient characteristics, and outcomes.

## 28. CONSENT

As per EUA requirements, physicians will discuss the risks and benefits of mABs and patients will consent to receive a mAB as part of usual care, should they desire mAB treatment. As per UPMC policy, the ordering physician reviews with patients the EUA Fact Sheet for each mAB and explain that the patient could receive any of the mABs governed by FDA EUAs.

## 29. ADVERSE EVENTS and SERIOUS ADVERSE EVENTS

The EUAs require providers and/or their designees report all medication errors and serious adverse events potentially related to the antibodies within seven calendar days from the onset of the event. Serious adverse events are defined as death, life-threatening event, inpatient hospitalization or prolongation of existing hospitalization, substantial disruption of ability to conduct normal life functions, a congenital anomaly/birth defect, or an intervention to prevent death, a life-threatening event, hospitalization, disability, or congenital anomaly.

The EUAs require adverse event reports be submitted to FDA MedWatch via one of multiple methods. Copies of all FDA MedWatch forms are also to be sent to the antibody manufacturer.

Thus, there already exist reporting requirements for UPMC associated with mAB prescription. We will track and record these reported data and adverse events by mAB assignment.

## 30. SAFETY and RISK MITIGATION

The EUAs stipulate warnings including hypersensitivity, clinical worsening, and side effects. As per EUA requirements, warnings will be communicated by providers to patients, adverse events will be reported as above, and post-infusion clinical monitoring will be done. Administration of mABs for patients with COVID-19 is routine care at UPMC, and their administration is not a research procedure.

## 31. MANAGEMENT of INFUSION REACTIONS

As per the EUAs, all participants should be monitored closely, as there is a risk of infusion reaction and hypersensitivity (including anaphylaxis) with any biological agent. Symptoms and signs that may occur as part of an infusion reaction include, but are not limited to fever, chills, nausea, headache, bronchospasm, hypotension, angioedema, throat irritation, rash including urticaria, pruritus, myalgia, and dizziness.

## 32. EXHIBITS

EUA fact sheets for health care providers

https://www.fda.gov/media/143603/download

https://www.fda.gov/media/143892/download

https://www.fda.gov/media/145802/download

## FINAL VERSION 1.3, June 30, 2021

UPMC OPTIMISE-C19 (<u>**OP**</u>timizing <u>**T**</u>reatment and <u>**I**</u>mpact of <u>**M**</u>onoclonal ant<u>**I**</u>bodie<u>**S**</u> through <u>**E**</u>valuation for COVID-19)

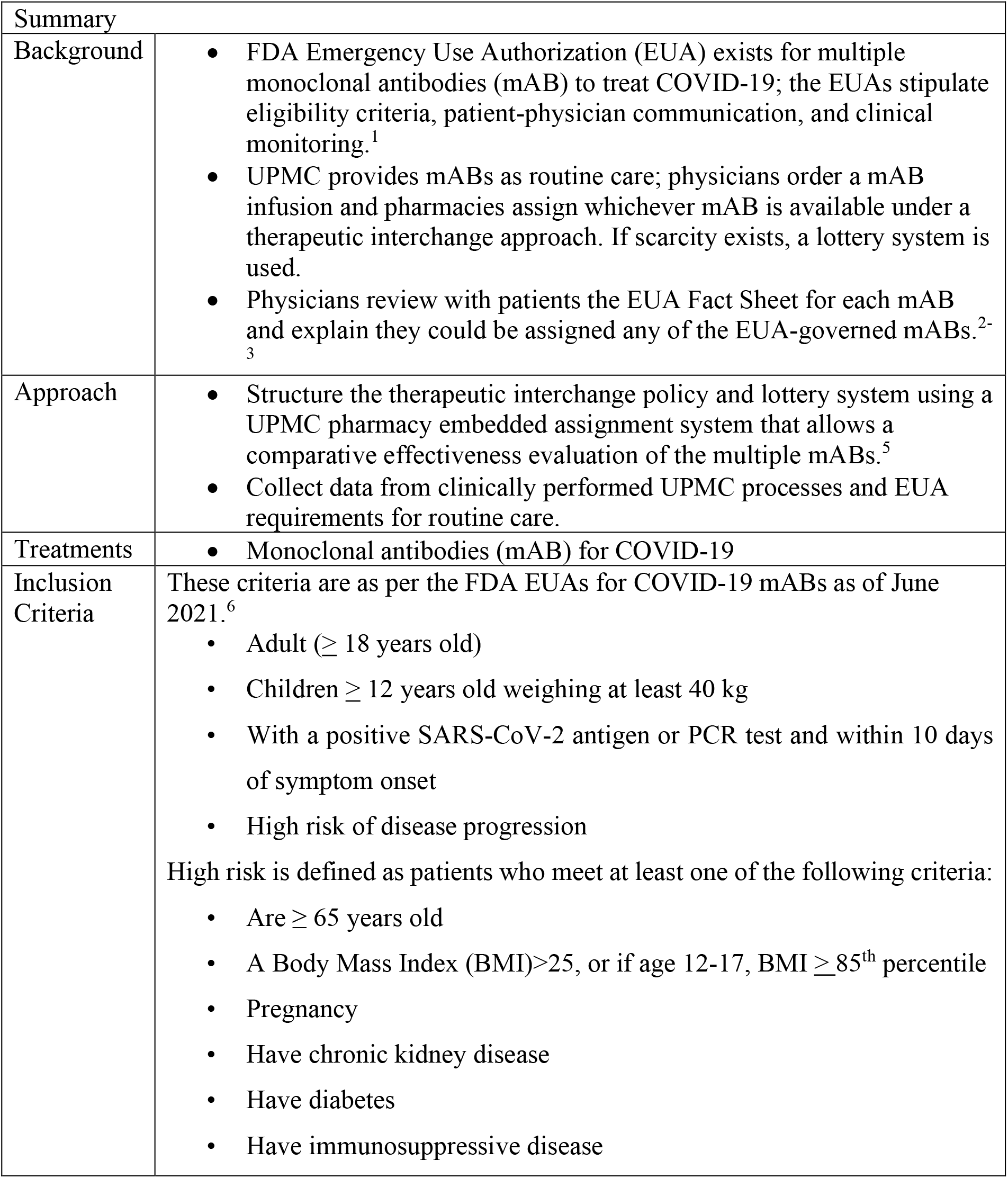
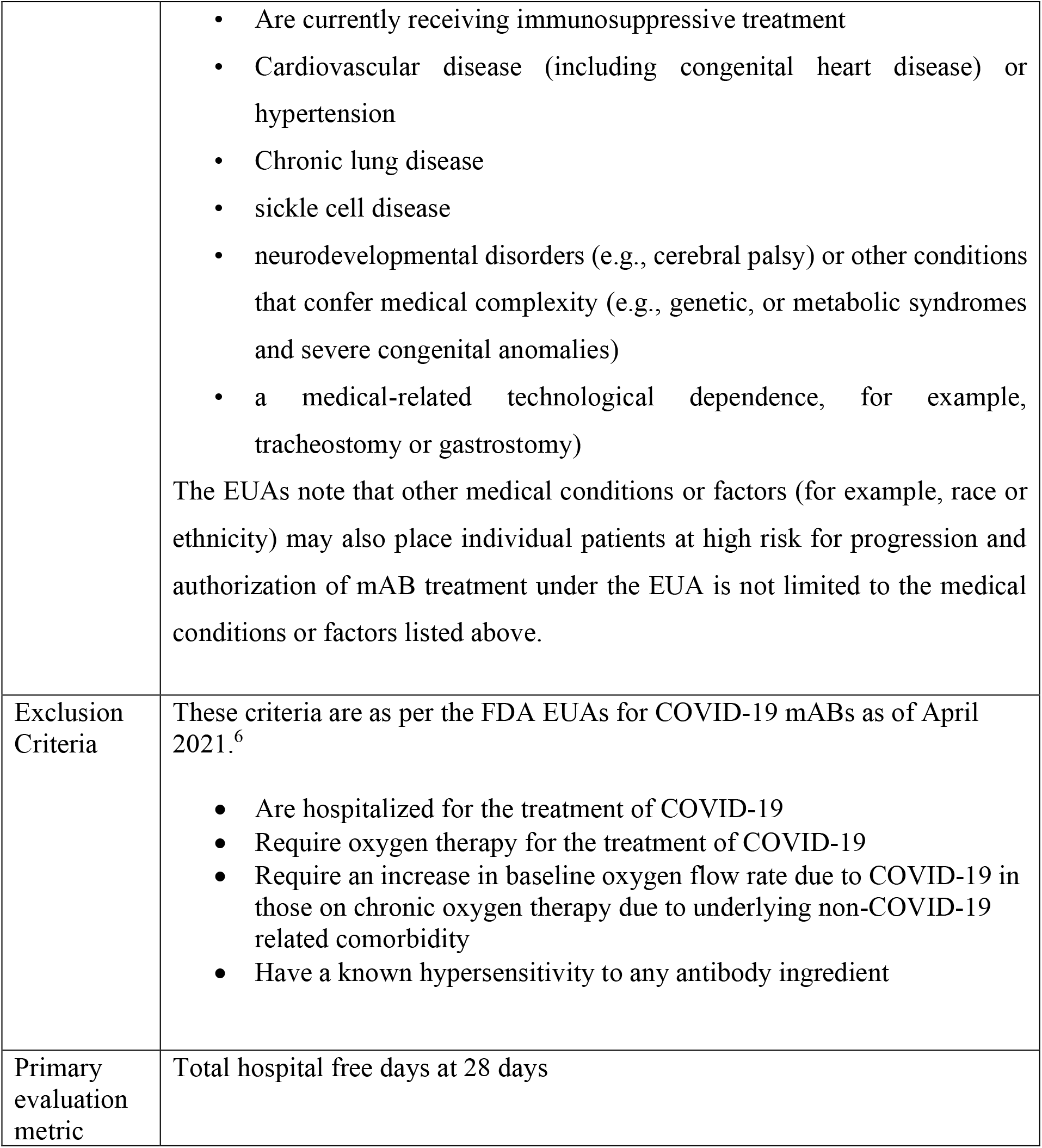

## 1. ABBREVIATIONS

AE: Adverse Events
BMI: Body Mass Index
C + I: casirivimab + imdevimab
CDC: Centers for Disease Control and Prevention
COPD: Chronic Obstructive Pulmonary Disease
COVID-19: Coronavirus disease 2019
CVD: Cardiovascular Disease
EUA: Emergency Use Authorization
FDA: Food and Drug Administration
HHS: Health and Human Services
HFD: Hospital Free Days
HTN: Hypertension
kDa: Kilodaltons
IgG1: Immunoglobulin G1
KG: Kilograms
OPTIMISE-C19: <u>**OP**</u>timizing <u>**T**</u>reatment and <u>**I**</u>mpact of <u>**M**</u>onoclonal ant<u>**I**</u>bodie<u>**S**</u> through <u>**E**</u>valuation
mAB: Monoclonal Antibodies
PCR: Polymerase Chain Reaction
SAEs: Serious Adverse Events
SARS-CoV-2: Severe Acute Respiratory Syndrome Coronavirus 2
UATRC: UPMC Antibody Treatment and Evaluation Center
Vir-7831: sotrovimab

## 2. BACKGROUND and RATIONALE

## 3. BACKGROUND

The SARS-CoV-2 mABs bamlanivimab and etesevimab, and the REGN-COV2 combination (casirivimab + imdevimab) reduce nasopharyngeal viral burden plus clinical outcomes including future emergency department visits and hospitalizations (Weinreich 33332778 NEJM, Gottlieb 33475701). Each received FDA Emergency Use Authorization (EUA) for use in selected populations (**Exhibit**); in April 2021 FDA revoked the EUA for bamlanivimab monotherapy, and in June 2021 FDA recommended bamlanivimab and etesevimab not be used. Additional trials of pre-exposure prophylaxis (NCT04497987) and other applications are underway, and additional mABs are in development.

In May 2021, it was announced that sotrovimab demonstrated clinical efficacy (85%) in reducing hospitalizations for more than 24 hours or death in those that received sotrovimab as compared to placebo (NCT04545060). Subsequently, it received EUA approval in select populations. Additional trials are underway.

This Appendix to the UPMC Pilot Core (PittPro 20040210) describes the approach of the UPMC OPTIMISE-C19 evaluation. We will conduct a pragmatic evaluation of monoclonal antibody treatments in participants with COVID-19 illness, starting with the patient population approved under the current FDA mAB EUAs.

## 4. RATIONALE

As of June 2021, there are over 10,000 new cases of COVID-19 diagnosed daily in the US https://covid.cdc.gov/covid-data-tracker/#trends_dailytrendscases, with over 1500 daily COVID-19 related hospital admissions Microsoft Power BI (powerbigov.us). Although case volumes are currently declining, COVID-19 remains a significant public health threat.

## 5. OBJECTIVES AND METRICS

## 9. POPULATION

We will evaluate patients that present to UPMC Emergency Departments, urgent care sites, infusions centers and other facilities that can or do provide mABs for COVID-19. As of June 30, 2021, there are 2 EUAs for COVID-19 mABs, with common inclusion and exclusion criteria, and we will evaluate patients that meet these criteria. As FDA antibody decisions change (E.g., FDA revokes or grants EUAs, or changes eligibility criteria), eligibility criteria will change.

## 10. INCLUSION CRITERIA

As per the current EUA criteria (June 2021), the following patients are included:

High risk is defined as patients who meet at least one of the following criteria:

- Are ≥ 65 years old
- A Body Mass Index (BMI)>25, or if age 12-17, BMI > 85^th^ percentile
- Pregnancy
- Have chronic kidney disease
- Have diabetes
- Have immunosuppressive disease
- Are currently receiving immunosuppressive treatment
- Cardiovascular disease (including congenital heart disease) or hypertension
- Chronic lung disease
- sickle cell disease
- neurodevelopmental disorders (e.g., cerebral palsy) or other conditions that confer medical complexity (e.g., genetic, or metabolic syndromes and severe congenital anomalies)
- a medical-related technological dependence, for example, tracheostomy or gastrostomy)

The EUAs note that other medical conditions or factors (for example, race or ethnicity) may also place individual patients at high risk for progression and authorization of mAB treatment under the EUA is not limited to the medical conditions or factors listed above.

## 11. EXCLUSION CRITERIA

As per the current EUA criteria (June 2021), the following are excluded:

## 12. EVALUATED TREATMENTS

We will evaluate mABs governed by FDA EUAs. Patients will receive COVID-19 mABs governed by FDA EUAs, when their treating physician orders a mAB and they meet EUA criteria. As FDA antibody decisions change (E.g., FDA revokes or grants EUAs, provides full approval, or changes eligibility criteria), available evaluated treatments will change. In April 2021, FDA revoked the EUA for bamlanivimab monotherapy and in June 2021 FDA recommended bamlanivimab and etesevimab not be used.

As of June 30, 2021, the EUA-approved mABs are as listed below.

## 14. SOTROVIMAB

Vir 7831 (sotrovimab) is a recombinant human IgG1k monoclonal antibody that binds to a conserved epitope on the spike protein receptor binding domain of SARS-CoV-2. Sotrovimab does not compete with human ACE2 receptor binding.

## 15. CONCOMITANT THERAPY

All care and concomitant therapy are as per the treating providers.

## 16. CONDUCT

## 17. DATA COLLECTION

- mAB was administered, including date, time, and infusion completion as well as the location of the infusion
- Demographics (including age, sex, race, body weight, vaccination status)
- Healthcare encounters, including hospital and ICU admission status if applicable
- Medication usage and doses
- Hospital and ICU admission status, if applicable
- Administration of medications related to COVID-19, if applicable
- Remnant blood availability
- Laboratory and microbiology data, including COVID-19 testing done for clinical purposes

## 19. ANTIBODY ADMINISTRATION

Antibodies will be administered as per the EUAs, UPMC Pharmacy and Therapeutics policies and the respective Pharmacy Manuals (as generated by the pharmaceutical companies), if applicable. Providers will explain mAB risks and benefits and provide the EUA Fact Sheets for Patients, Parents and Caregivers as per EUA requirements.

## 21. STATISTICAL CONSIDERATIONS

## 23. NUMBER of PARTICIPANTS

Sample size is determined by case volume throughout the course of the pandemic.

## 25. ETHICAL CONSIDERATIONS

## 26. DATA MONITORING

UPMC clinical leadership will regularly monitor monthly reports on enrollment, patient characteristics, and outcomes. Leadership will also receive regular interim analyses from the adaptive statistical model to inform UPMC clinical policy.

## 31. EXHIBITS

## EUA fact sheets for health care providers

https://www.fda.gov/media/149534/download (sotrovimab) https://www.regeneron.com/downloads/treatment-covid19-eua-fact-sheet-for-hcp.pdf (casirivimab and imdevimab)

## SUMMARY OF CHANGES

OPTIMISE-C19 - Summary of Changes for Amendment 1 on April 25, 2021:

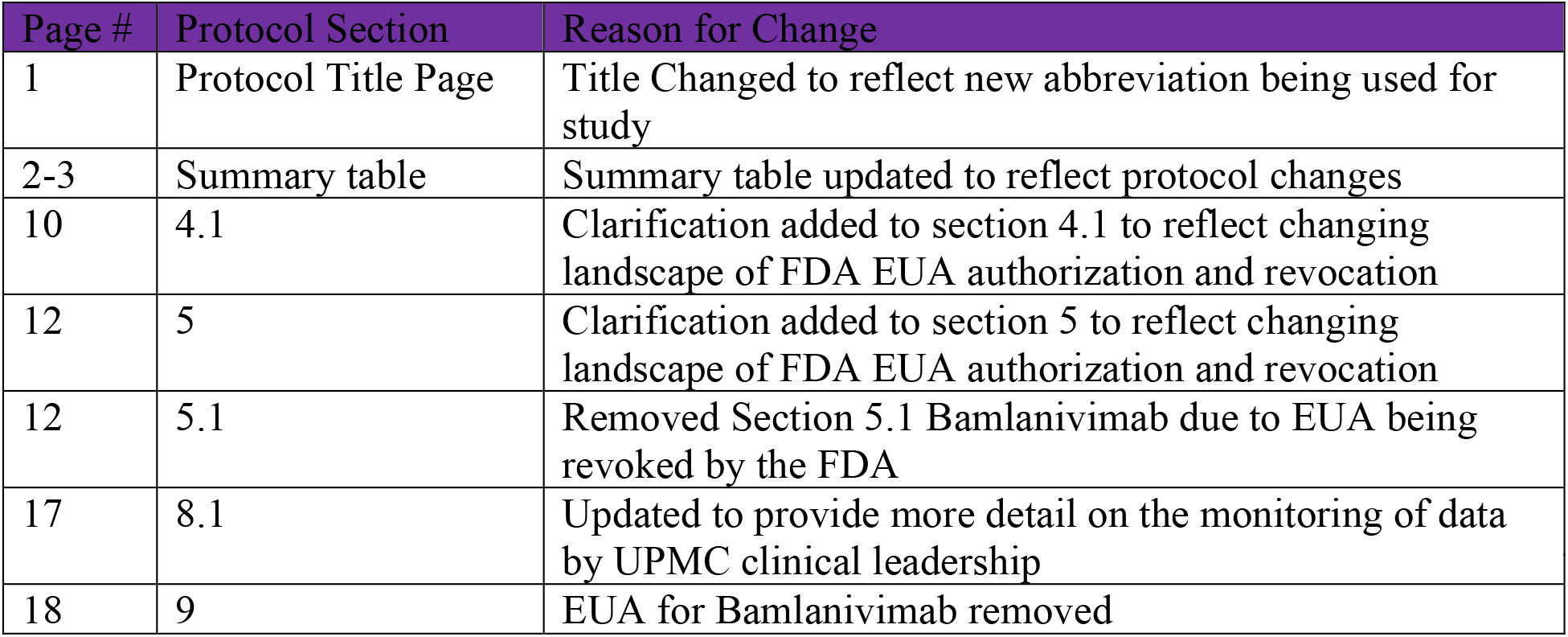

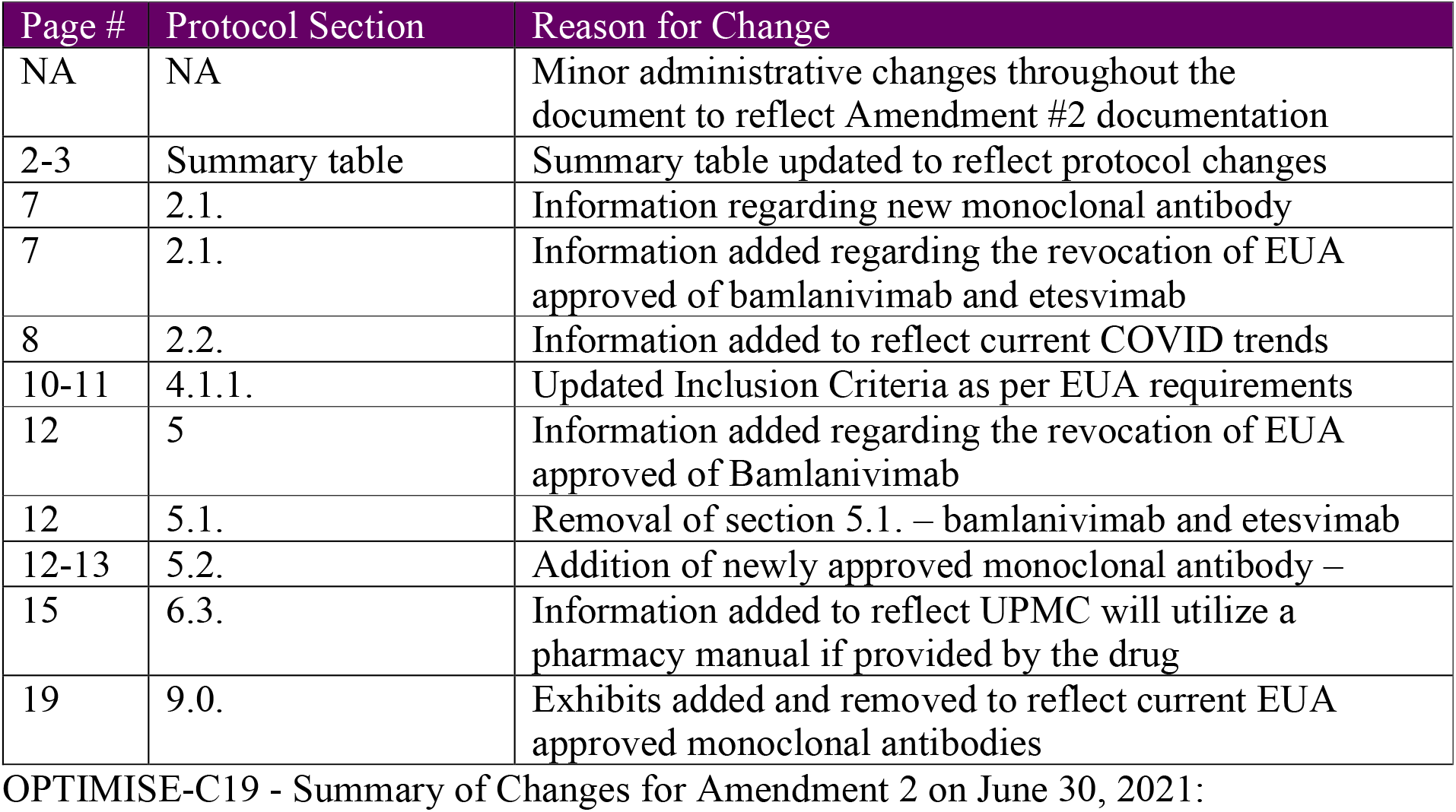

## STATISTICAL ANALYSIS PLAN

### 1. Introduction

This document describes the detail of the analysis read-out from July 26, 2021. This document is an appendix to the Statistical Analysis Plan for the “UPMC Antibody Treatment and Evaluation Center” with the details for this analysis read-out.

- We will analyze and report three treatment arms (below) as the first two have been administratively closed to enrollment due to FDA decisions.
- Unblinded data from this and prior interim analyses can be shared with investigators after the last randomized allocations (June 25) to the two mAB treatment arms closed by FDA.
- This unblinding is appropriate as future analyses of patients randomized to the third arm (C+I) will not be compared to the first two arms, there are no “control” arms as all patients receive mAB treatment, and future comparisons will be of C+I vs newer mABs.
- Enrollment continues in the currently available treatment arms (C+I and S). “S” refers to sotrovimab, produced by GSK and Vir.

### 2. Treatment Arms

There are three treatment arms that will be included in this analysis. The treatment arms are

1. B (Bamlanivimab)
2. B+E (Bamlaniviman/Etesevimab combination)
3. C+I (casirivimab/imdevimab combination)

### 3. Primary Endpoint

The primary endpoint for this read-out is hospital-free days.

### 4. Primary Analysis Population

The primary analysis population for this read-out is the “As-Infused” population. This analysis will include each patient randomized from March 10, 2021 until June 25, 2021. The date of the data set snapshot is on July 26, 2021.

### 5. Primary Analysis Model

The primary analysis model is as described in the trial SAP.

The primary analysis for the primary endpoint is a cumulative proportional odds model. Let the probability of an outcome of less than or equal to y be *π*_*y*_ = *Pr*(*Y* ≤ *y*). Let *a* be the indicator of treatment arm (*a*=1,…,k). The model adjusts for the following baseline variables:

1. ED or infusion center (0=infusion center, 1=ED)
2. Age (with categories of <30, 30-39, 40-49, 50-59, 60-69, 70-79, and ≥80; 60-69 will be used as the referent)
3. Sex (sex at birth, male is the referent)
4. Covid variant is not modeled in this primary analysis
5. Time (two-week epochs of time are used for adjustments; the most current 4-week period is the referent)

The primary analysis model is based on a cumulative logistic regression, where *π*_*y*_ = *Pr*(*Y* ≤ *y*), where

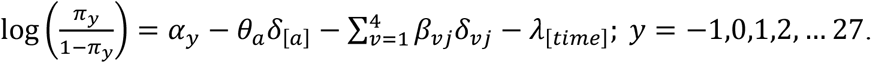

The additive covariate effects across all treatment arms for each patient are modeled through the *β* parameters. The *δ* parameters are indicator functions for the treatment arm and covariate values for the baseline covariates. The efficacy of the treatment arms is modeled with the *θ* parameters. The ordinal effect parameters (*α*_*y*_) are modeled with a Dirichlet distribution with equal weight on each outcome and a sum of 1.

The baseline covariate effects are modeled with independent weak prior distributions:

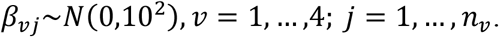

The appropriate coefficients will be set to 0 within each covariate for identifiability (the goal will be the largest category set to 0).

The effects of time are adjusted within the model using two-week epochs and a smoothing model over time. The modeling of the time effects is set up with the most current period (2 epochs combined being the most recent month are set to 0):

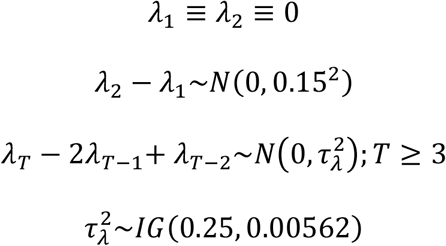

The prior distributions for the mAB treatment effects are weak:

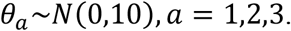

### 6. The treatment arms with the largest sample size should be selected as the referent arm (label a=1)_for treatment effects and assigned a treatment effect of *θ*_1_ = 0

### 7. Model Convergence

Given the complexity of the model, conventions may be taken by the analysis team if there are convergence issues or model stability issues. For example, there may be outcome categories in the 30 possible primary outcome values (e.g., k number of hospital-free days, patient death) that do not occur. If this happens at analysis, the cells will be combined to achieve model convergence. For example, if the 4 hospital-free day outcome value does not occur it will be combined with 3, and so on, until every cell has occurred. Additional model stability conventions will be taken to preserve the model stability.

### 8. Missing and Partial Data

If there are missing covariates for a patient in the as-infused patient population, the following conventions will be used.

1. If the treatment arm is missing the patient will be ignored.
2. If a baseline covariate is missing the referent value for that covariate will be used

For all model analyses, only patients who have achieved 28-days of follow-up from the date of the index infusion will be used in the analysis. No use or imputation of patient data for patients with less than 28 days will be conducted.

Given the HER-based data summaries there will be no missing outcome data. If there is deemed to be a corrupted outcome that patient will be ignored. Some patients may have 28 hospitalization-free days that at subsequent analyses are found to have out of system hospitalizations. The data will be updated at future analyses.

### 9. Trial Inferences

For the primary analysis, there is no “control” treatment and so all inferences are made comparing the individual treatment arms to each other. The main quantity of interest will be the relative odds ratio between any two treatments arms

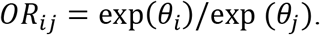

The posterior probability that the odds ratio for arm *i* compared to arm *j* is greater than 1 (signifying that treatment *i* is superior to treatment *j*) is used as a comparison between arms. Additionally, the posterior mean and 95% confidence interval between arms will be used to summarize relative treatment effects.

#### Arm Inferiority

If one of the arms has a 99% chance of being inferior to any of the other available arms then the inferior arm will be declared inferior and may be removed from the trial. There may be conditions of the pandemic (variation frequency, new variations) or drug supply concerns that an arm is retained.

#### Equivalence

Any two arms in the trial may reach a declaration of equivalence. It is anticipated that no actions would take when equivalence is reached but a declaration and public disclosure may be made. There is a sliding scale of equivalence with different levels of equivalence bounds.

A declaration of equivalence will be tied to the equivalence level. Equivalence with a bound of d is declared if the posterior probability of the odds ratio is with d is at least 95%:

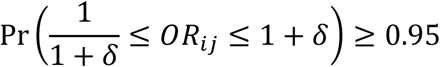

The following levels are pre-defined:

1. The first level of equivalence occurs when there is 95% posterior probability that the odds-ratio is within a bound of d=0.25
2. The second level of equivalence occurs when there is a 95% posterior probability that the odds-ratio is within a bound of d=0.20
3. The third level of equivalence occurs when there is a 95% posterior probability that the odds-ratio is within a bound of d=0.15
4. The fourth level of equivalence occurs when there is a 95% posterior probability that the odds-ratio is within a bound of d=0.10
5. The fifth level of equivalence occurs when there is a 95% posterior probability that the odds-ratio is within a bound of d=0.05

#### Combination Futility

For comparing B+E to B the combination (B+E) will be compared to the individual component arm (B). If there is more than a 95% probability that the effect of the combination B+E is no better than a 20% improvement in the odds ratio compared to B, then the combination will be declared *not clinically relevantly superior (combination futile)* to N.

### 10. Modeling Treatment Heterogeneity Across Variant Date Prevalence Epochs

For this read out, the following time epochs will be modeled with different treatment effects using the Treatment Heterogeneity analysis model.

The treatment effect, *θ*_*a*_, will be modeled separately within each epoch, *s* = 1, …, *S*, as *θ*_*a*1_, *θ*_*a*2_, …, *θ*_*aS*_, with hierarchical prior distribution:

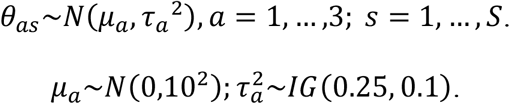

The following time epochs are specified:

1. March 10 - March 31
2. April 1 – April 30
3. May 1 – May 31
4. June 1 – June 25

Specific Analyses

| # | Status | Population | Endpoint | Other |
| --- | --- | --- | --- | --- |
| **1** | Primary | As-Infused to B, B+I, or C+I on or before June 25, 2021 | HFD |  |
| **2** | Variant Secondary | As-Infused to B, B+I, or C+I on or before June 25, 2021 | HFD | Differential efficacy by time epoch specified |

The primary analysis

The following posterior probabilities will be reported

The primary analysis The following posterior probabilities will be reported
| **Quantity of Interest** | **Posterior Probability** |
| --- | --- |
| B+E superior to B |  |
| B+E combination futile to B |  |
| C+I superior to B |  |
| C+I superior to B+E |  |

Statistical Triggers Met for Equivalence with delta ranges of equivalence.

The following will be reported:

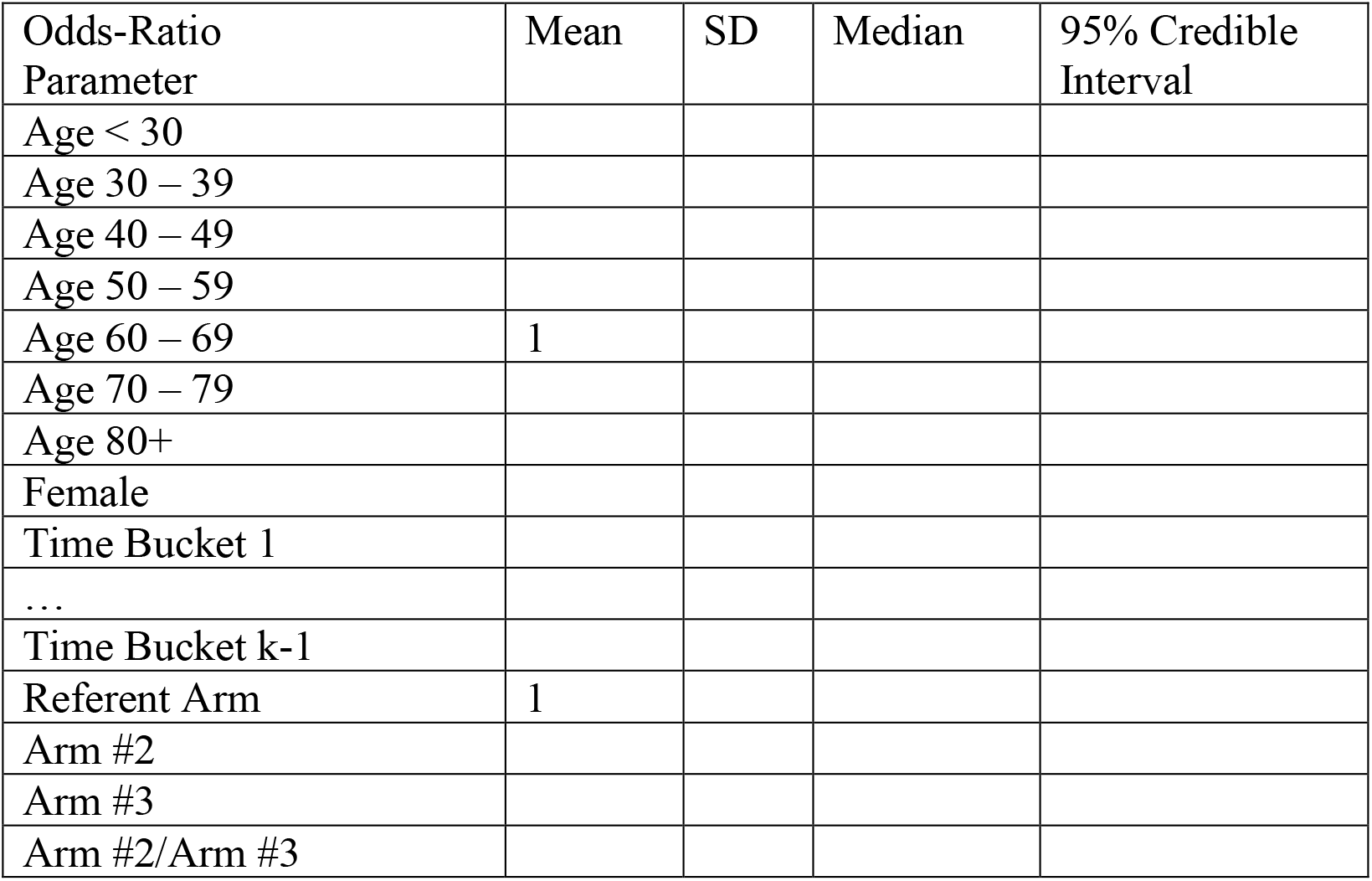

Statistical Triggers Met for Equivalence with delta ranges of equivalence. The following will be reported:
| Odds-Ratio Parameter | Mean | SD | Median | 95% Credible Interval |
| --- | --- | --- | --- | --- |
| Age < 30 |  |  |  |  |
| Age 30 – 39 |  |  |  |  |
| Age 40 – 49 |  |  |  |  |
| Age 50 – 59 |  |  |  |  |
| Age 60 – 69 | 1 |  |  |  |
| Age 70 – 79 |  |  |  |  |
| Age 80+ |  |  |  |  |
| Female |  |  |  |  |
| Time Bucket 1 |  |  |  |  |
| ... |  |  |  |  |
| Time Bucket k-1 |  |  |  |  |
| Referent Arm | 1 |  |  |  |
| Arm #2 |  |  |  |  |
| Arm #3 |  |  |  |  |
| Arm #2/Arm #3 |  |  |  |  |

Graphical summaries

1. Stacked bar plots and cumulative distributions of HFDs by treatment arm
2. Stacked bar plots and cumulative distributions of HFDs by time epochs
3. Stacked bar plots and cumulative distributions of HFDs by sex

1. The Variant Secondary analysis

The following posterior probabilities will be reported for each time epoch

| Quantity of Interest | Posterior Probability |
| --- | --- |
| B+E superior to B |  |
| B+E combination futile to B |  |
| C+I superior to B |  |
| C+I superior to B+E |  |

Statistical Triggers Met for Equivalence with delta ranges of equivalence

The following will be reported:

Statistical Triggers Met for Equivalence with delta ranges of equivalence The following will be reported:
| Odds-Ratio<br>Parameter | Mean | SD | Median | 95% Credible<br>Interval |
| --- | --- | --- | --- | --- |
| Age < 30 |  |  |  |  |
| Age 30 – 39 |  |  |  |  |
| Age 40 – 49 |  |  |  |  |
| Age 50 – 59 |  |  |  |  |
| Age 60 – 69 | 1 |  |  |  |
| Age 70 – 79 |  |  |  |  |
| Age 80+ |  |  |  |  |
| Female |  |  |  |  |
| Time Bucket 1 |  |  |  |  |
| ... |  |  |  |  |
| Time Bucket k-1 |  |  |  |  |
| Referent Arm | 1 |  |  |  |
| For each time-epoch |  |  |  |  |
| Arm #2 |  |  |  |  |
| Arm #3 |  |  |  |  |
| Arm #2/Arm #3 |  |  |  |  |

Graphical summaries

4. Stacked bar plots and cumulative distributions of HFDs by treatment arm by time epoch

## Standard CONSORT with Pragmatic Trials Checklist

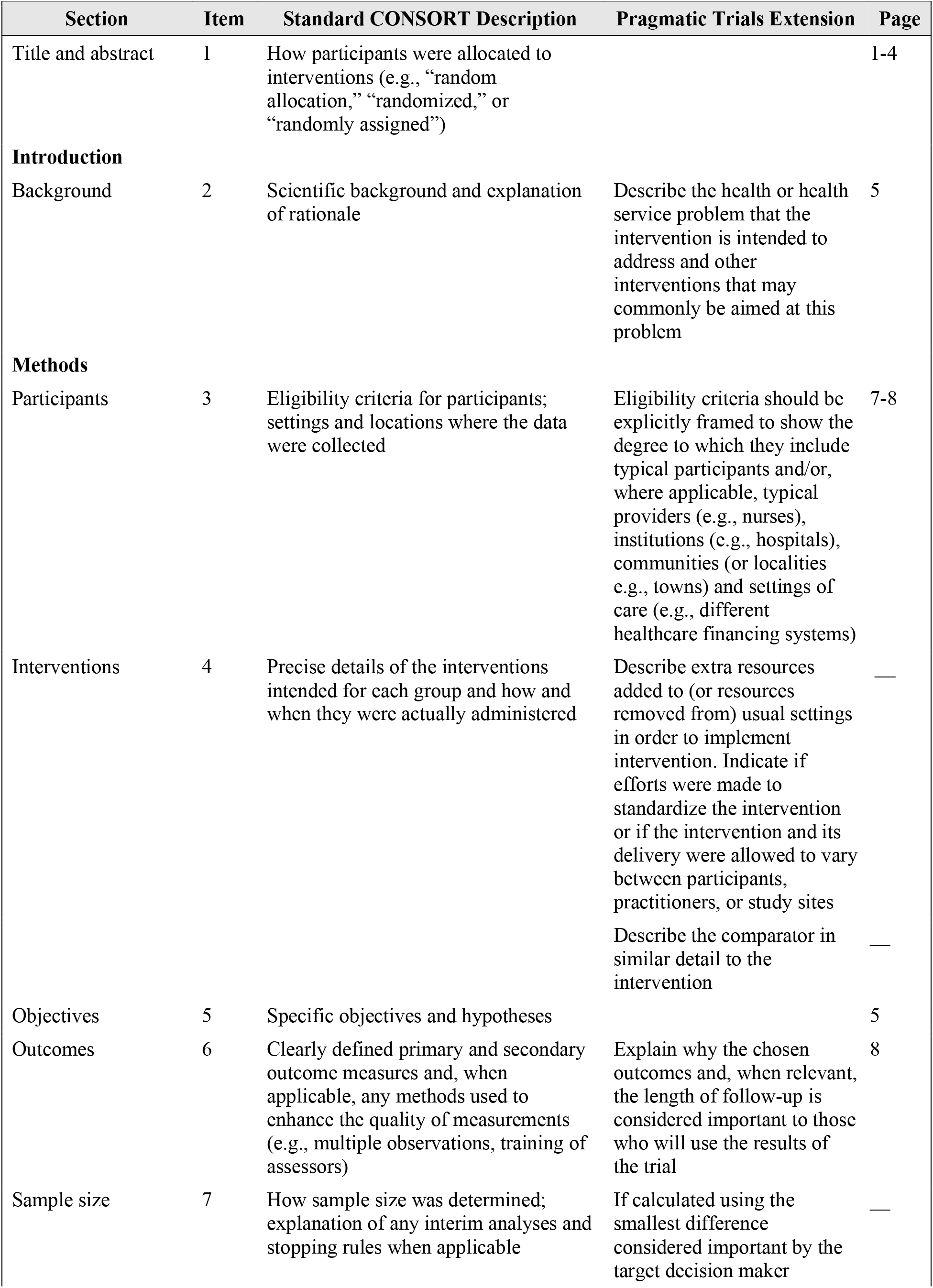
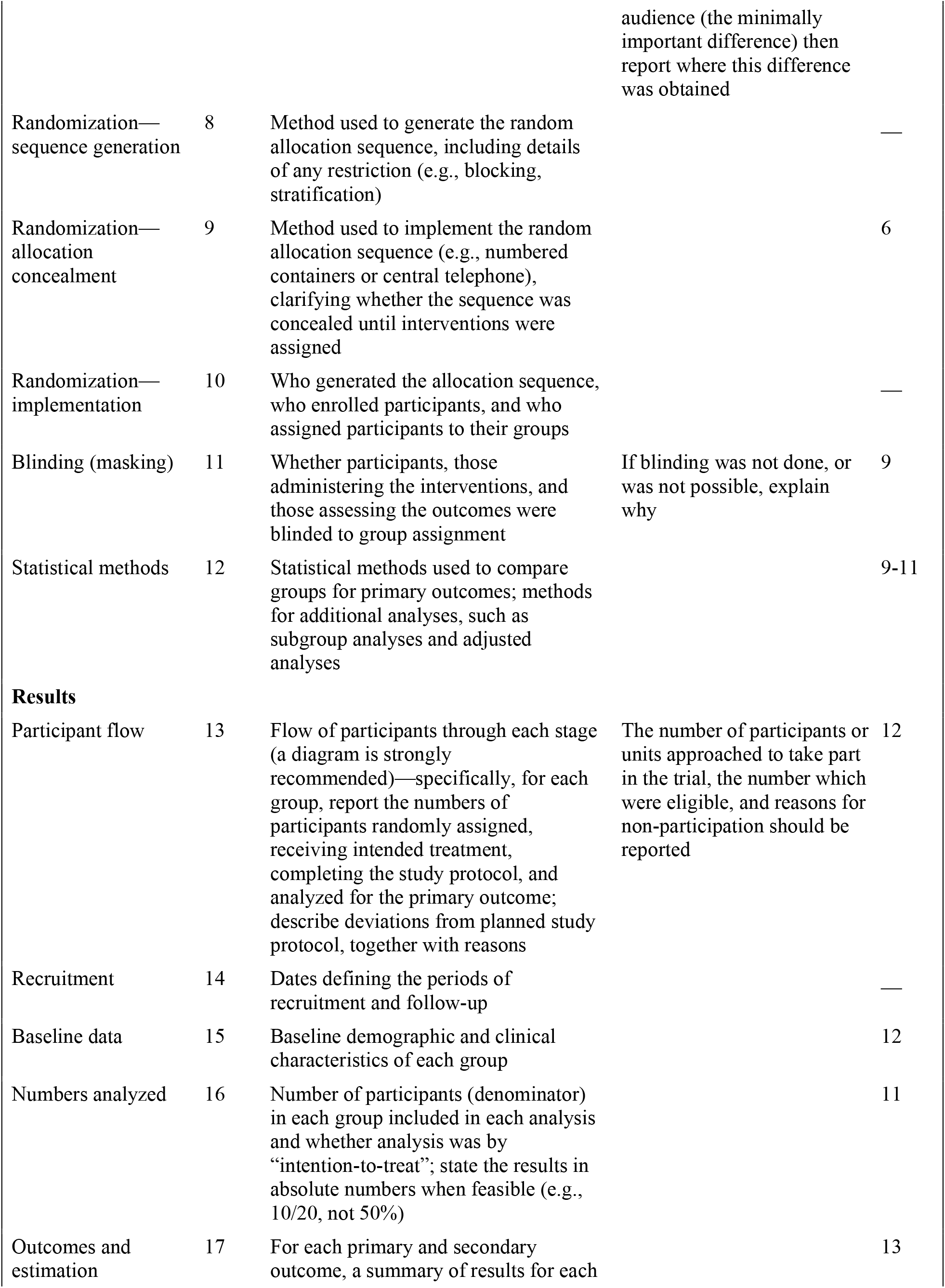
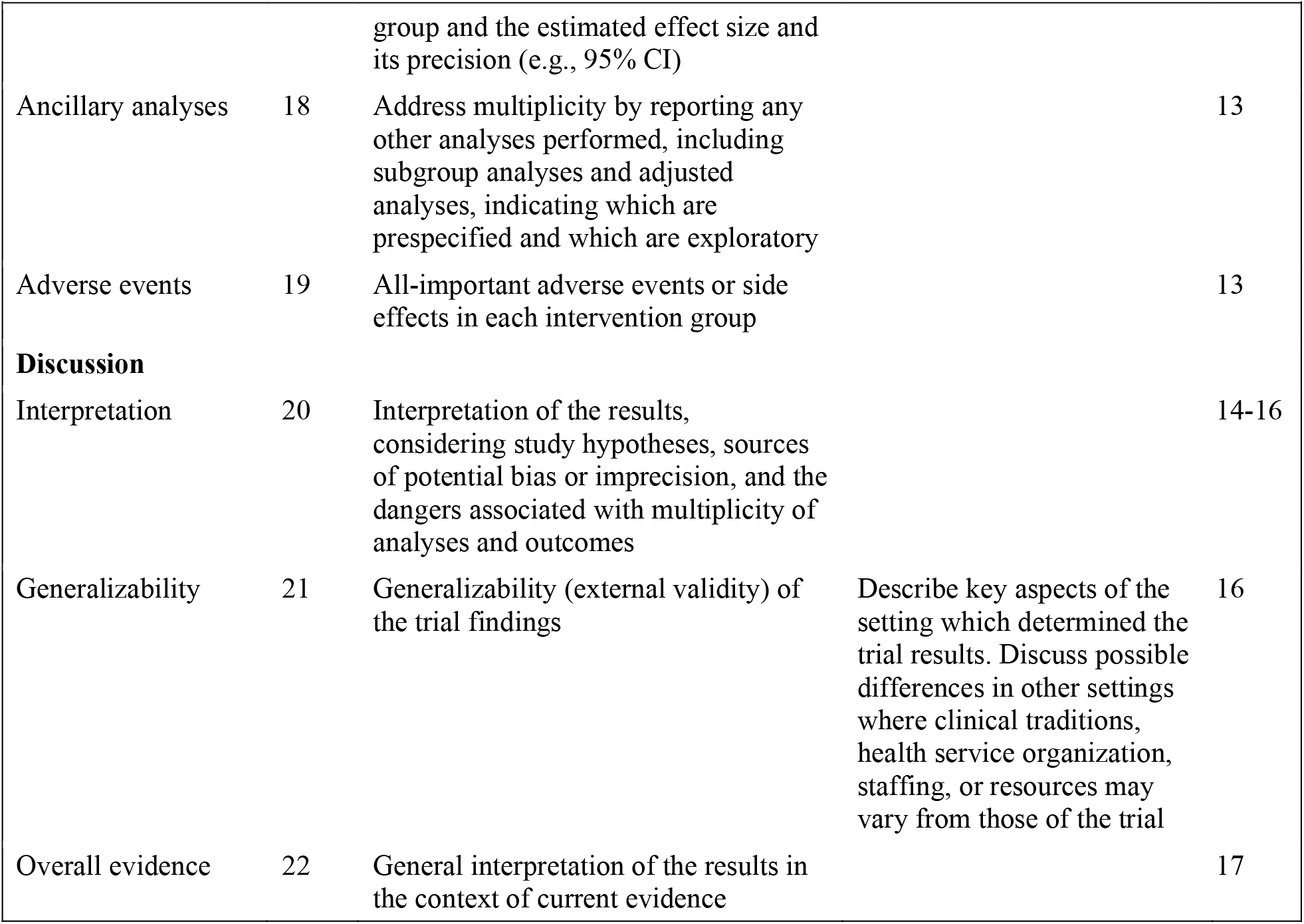

